# Why Daily SARS-CoV-2 Nasal Rapid Antigen Testing Poorly Detects Infected and Infectious Individuals

**DOI:** 10.1101/2022.07.13.22277513

**Authors:** Alexander Viloria Winnett, Reid Akana, Natasha Shelby, Hannah Davich, Saharai Caldera, Taikun Yamada, John Raymond B. Reyna, Anna E. Romano, Alyssa M. Carter, Mi Kyung Kim, Matt Thomson, Colten Tognazzini, Matthew Feaster, Ying-Ying Goh, Yap Ching Chew, Rustem F. Ismagilov

## Abstract

**Background:** In a recent household-transmission study of SARS-CoV-2, we found extreme differences in SARS-CoV-2 viral loads among paired saliva, anterior-nares swab (ANS) and oropharyngeal swab specimens collected from the same timepoint. We hypothesized these differences may hinder low-analytical-sensitivity assays (including antigen rapid diagnostic tests, Ag-RDTs) using a single specimen type (e.g., ANS) from reliably detecting infected and infectious individuals.

**Methods:** We evaluated a daily at-home ANS Ag-RDT (Quidel QuickVue) in a cross-sectional analysis of 228 individuals and in a longitudinal analysis (throughout infection) of 17 individuals enrolled early in the course of infection. Ag-RDT results were compared to RT-qPCR results and high, presumably infectious viral loads (in each, or any, specimen type).

**Results:** The ANS Ag-RDT correctly detected only 44% of timepoints from infected individuals on cross-sectional analysis, and in this population had an inferred limit of detection of 7.6×10^6^ copies/mL. From the longitudinal cohort, daily Ag-RDT clinical sensitivity was very low (<3%) during the early, pre-infectious period of the infection. Further, the Ag-RDT detected ≤63% of presumably infectious timepoints. The poor observed clinical sensitivity of the Ag-RDT was similar to what was predicted based on quantitative ANS viral loads and the inferred limit of detection of the ANS Ag-RDT being evaluated, indicating high-quality self-sampling.

**Conclusion:** Nasal Ag-RDTs, even when used daily, can miss individuals infected with the Omicron variant and even those presumably infectious. Evaluations of Ag-RDT detection of infected or infectious individuals should be compared with a composite (multi-specimen) infection status to correctly assess performance.

**Key points:** Nasal-swab rapid antigen tests have low analytical sensitivity and the sampling of only the nasal cavity hinders their ability to detect infected individuals, including those with high and presumably infectious viral loads in throat or saliva specimens.

## BACKGROUND

Earliest detection of SARS-CoV-2 infections can substantially reduce transmission, minimize the spread of new variants, and prompt sooner treatment initiation for better patient outcomes. Antigen rapid diagnostic tests (Ag-RDTs) with nasal swabs are increasingly used for SARS-CoV-2 screening and diagnosis globally.^1-6^ Indeed, Ag-RDTs are a powerful tool given the low cost (compared with molecular tests), speed, and portability—all of which improve accessibility in remote or lower-resourced settings and at-home use.^5,7,89^ However, Ag-RDTs and some rapid molecular tests have lower analytical sensitivity than most gold-standard reverse-transcription quantitative PCR (RT-qPCR) tests and therefore require high viral loads (typically greater than 10^5^ copies/mL) to reliably yield positive results.^7,8,10-18^ Further, Ag-RDTs are frequently used in unintended ways.^15,19^ For example, although many Ag-RDTs are not authorized for asymptomatic use and/or have poor clinical sensitivity in asymptomatic populations^20-23^ they are widely used for test-to-enter and serial-screening purposes.^14,21,24^

One view of Ag-RDTs is that these tests (and molecular tests with low analytical sensitivity) can still prevent or mitigate SARS-CoV-2 outbreaks if they are used frequently.^25-27^ This view is based on the assumption that Ag-RDTs may miss some infected individuals, but will result positive when individuals are infectious with high viral loads detectable by these low-analytical-sensitivity assays.^27-29^ This concordance would allow high-frequency Ag-RDTs (with immediate results) to more effectively prompt isolation of infectious individuals than a high-analytical-sensitivity test (with delayed results).^26,27^

Investigating the performance of Ag-RDTs for detecting the infectious period is challenging, because assessing replication-competent virus in clinical specimens complicated.^29-31^ Viral culture is difficult, costly, and restricted to specialized laboratories. Thus, direct investigations of infectious virus are infrequently performed. Instead, because replication-competent virus is associated with specimens with high viral loads (10^4^ copies/mL and above) in studies that have performed SARS-CoV-2 viral culture,^16,25,30-47 29^ viral load is often used as a surrogate for infectiousness.

Data in longitudinal studies that captured viral load measurements from early in the course of infection^31,41,43,47-58^ present evidence that challenge the assumption that Ag-RDTs will reliably detect pre-infectious and infectious individuals. Viral-load timecourses from individuals in these studies show that for some individuals, several days can pass between when viral loads reach potentially infectious levels and when viral loads rise to the limits of detection (LODs) of Ag-RDTs (approximately 10^5^ to 10^7^ copies/mL).^7,8,10-17,51,52^. During this window, previously conceptualized as a period of cryptic transmission,^59^ false-negative Ag-RDT results may embolden social contact and increase transmission.^60,61^

Additionally, in our recent large household-transmission study analyzing viral loads from daily sampling of anterior-nares nasal swabs (ANS), oropharyngeal swabs (OPS) and saliva (SA) beginning from the incidence of SARS-CoV-2 Omicron infection, two major findings emerged that suggest Ag-RDTs may miss many pre-infectious and infectious individuals.^55^ First, we found that viral loads differed significantly (by more than 9 orders of magnitude) among different specimen types from the same person at the same timepoint, and that viral load timecourses in each specimen type from a person did not correlate with each other over time. Individuals often had high and presumably infectious viral loads in one specimen type (e.g., OPS), yet very low viral loads in another type (e.g., ANS). Because all at-home Ag-RDTs authorized by the U.S. Food and Drug Administration (FDA) are authorized only for use with nasal swabs,^62^ the lack of correlation among viral loads in different specimen types could hinder the ability of nasal swab Ag-RDTs to detect infectious individuals who have high viral loads in specimen types other than nasal swab. Second, we observed that most individuals exhibit a delay in the rise of ANS viral loads relative to specimens from the oral cavity^55^; this finding is consistent with previous reports by us^52^ for ancestral SARS-CoV-2 variants and other studies^41,43,49,51^ that included the early period of infection in multiple specimen types. A delay in the rise of ANS viral loads could delay detection of infected and infectious individuals by nasal swab Ag-RDTs.

Although many Ag-RDT evaluations have observed relatively high concordance between Ag-RDT results and infectiousness (as established directly by viral culture^21,34,41,46,48-50,63-70^ or presumed by quantitative viral loads or semi-quantitative Ct values^22,71-73^), this high concordance may not hold when Ag-RDTs are used for screening testing in the community. For example, in several studies^34,46,48,64,68,70-73^ most infected participants were already symptomatic by the time sampling began, so Ag-RDT performance would not be generalizable to the early period of infection. A few studies accounted for infection stage and assessed longitudinal performance of nasal swab Ag-RDTs relative to viral culture^41,49,50,63,65,66,70^, although only some^41,49,50,63,65^ were designed for prospective sampling that would capture the early period of infection. Importantly, none of these studies tested for infectious virus in multiple upper-respiratory specimen types. To our knowledge, only one study evaluating a nasal swab Ag-RDT examined infectiousness in an oral specimen type; this study found that an ANS Ag-RDT was often negative while individuals had infectious viral loads in saliva.^51^

Here, we report on a field evaluation of an ANS Ag-RDT (the Quidel QuickVue At-Home OTC COVID-19 Test), with both cross-sectional and longitudinal analyses. The ANS Ag-RDT was taken prospectively each day by participants with a recently infected or exposed household contact. Participants also collected daily SA, ANS, and OPS specimens for SARS-CoV-2 testing and viral-load quantification by high-analytical-sensitivity RT-qPCR tests, as previously described.^55^ From these viral-load measurements we assessed performance of the ANS Ag-RDT to detect individuals with detectable SARS-CoV-2 in any of the three specimen types tested, as well as those with presumably infectious viral loads in any of the three specimen types. This design allowed us to probe not only the performance of this Ag-RDT for early detection of infected and infectious individuals, but also potential reasons why many Ag-RDTs may exhibit poor performance to detect infected and infectious individuals.

## METHODS

### Study Design

We performed a case-ascertained study in the greater Los Angeles County area between November 2021 and March 2022 in which participants completed a daily online symptom survey, then prospectively self-collected saliva (SA), then anterior-nares (nasal) swab (ANS), then posterior oropharyngeal (throat) swab (OPS) specimens for high-analytical-sensitivity RT-qPCR testing. Details of RT-qPCR sampling and the results of the symptoms analysis are provided in a separate report.^55^ After self-collecting these specimens, participants immediately performed an at-home ANS Ag-RDT (the FDA-authorized Quidel QuickVue At-Home OTC COVID-19 Test^74^) per manufacturer instructions^75^ and reported the result and submitted a photograph of the test strip via a secure REDCap server. This Ag-RDT is authorized and in use globally,^2^ with performance for ancestral variants evaluated in several cross-sectional studies.^10,23,76,77^

The manufacturer reports that this Ag-RDT has an LOD with >95% positivity at 1.91×10^4^ TCID50/mL of commercial heat-inactivated SARS-CoV-2 particles.^74^ Conversion of TCID50/mL to viral load in copies/mL is not provided in the FDA documentation, and the manufacturer was unable to provide this value nor a lot number or certificate of analysis for the heat-inactivated particles. Thus, we were unable to convert this LOD value from TCID50/mL to copies/mL.

The RT-qPCR data (test result, and viral-load quantifications) were compared to Ag-RDT results for both cross-sectional and longitudinal analyses of Ag-RDT performance (**Fig 1**). The 228 participants who enrolled in the study (**Table S1**) provided data for 2,215 timepoints for cross-sectional analysis. A composite RT-qPCR result was generated for each of the timepoints: a participant was considered infected if any of their three specimen types yielded a positive result by RT-qPCR; a participant was considered uninfected if all specimen types resulted negative by RT-qPCR; and results were inconclusive if at least one specimen type resulted inconclusive while the other specimen types resulted negative by RT-qPCR. A total of 2,174 timepoints specimens had a valid, conclusive composite RT-qPCR result, of which 847 timepoints from 90 individuals were classified as infected. Of these 2,174 timepoints, 63 did not have associated Ag-RDT results reported by the participant and 4 had invalid results. Three positive Ag-RDT results were also excluded because they originated from a faulty lot of test strips (see Supplementary Information). A total of 2,107 (nasal swab), 2,108 (throat swab), and 2,114 (saliva) timepoints had valid ANS Ag-RDT and RT-qPCR results (**Fig 2A-F, Fig 3A-C**), and 2,104 timepoints had valid, paired ANS Ag-RDT and composite RT-qPCR results (**Fig 2G-H, Fig 3D-F**). For analyses oriented to early infection (**Fig 4, Fig 5, Fig 6**), we analyzed longitudinal data from 17 participants (**Fig 1A, Table S2**) who were initially negative in at least one test (either RT-qPCR or Ag-RDT) upon enrollment, indicating they enrolled early in the course of the infection.

**Figure 1.**
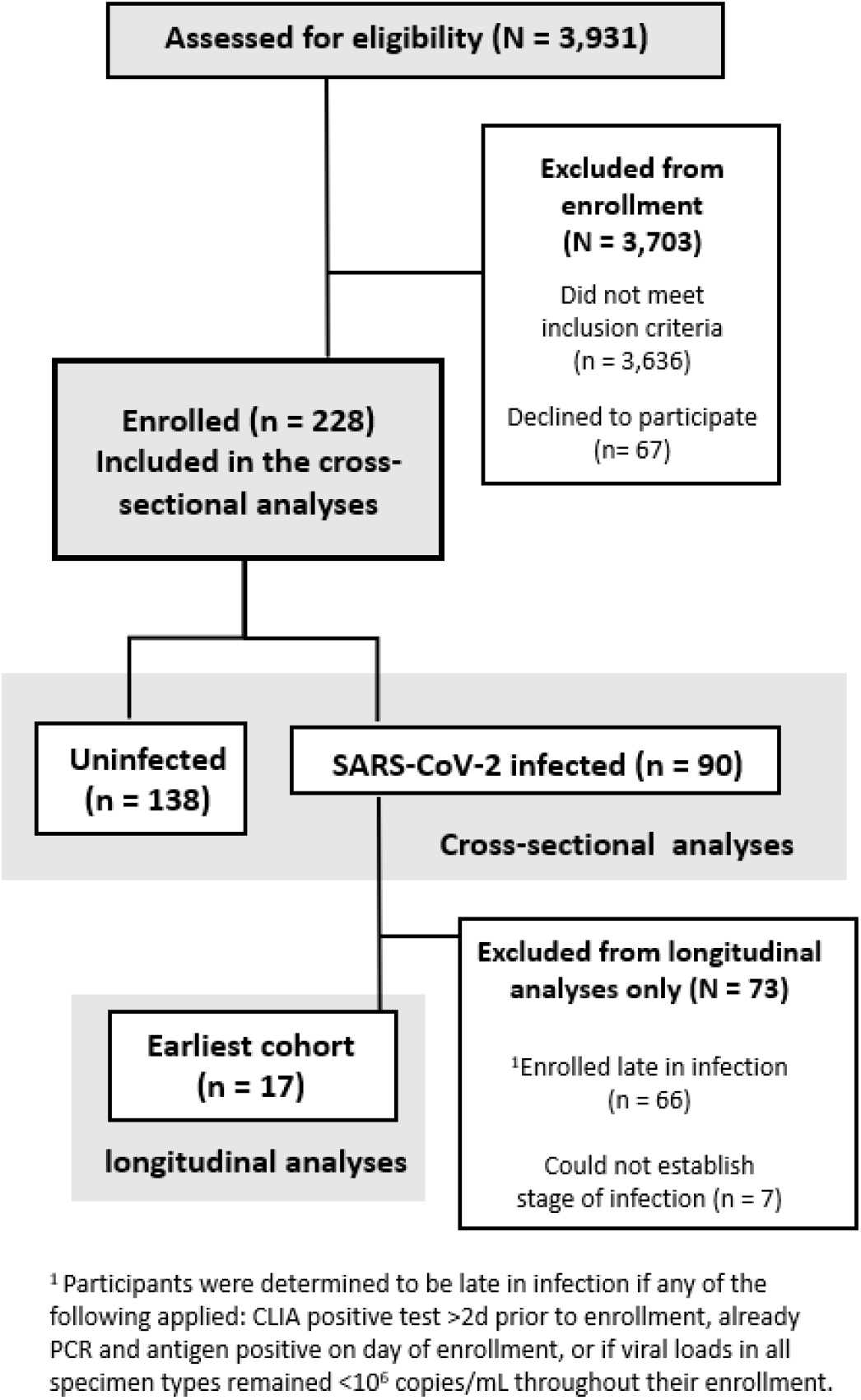
A CONSORT diagram of participant recruitment, eligibility, and enrollment for the cross-sectional and longitudinal analyses. Demographic and medical information can be found in **Tables S1-S2**. Cross-sectional analyses are presented in **Fig 2-3**; longitudinal analyses are presented in **Fig 4-6**.

**Figure 2.**
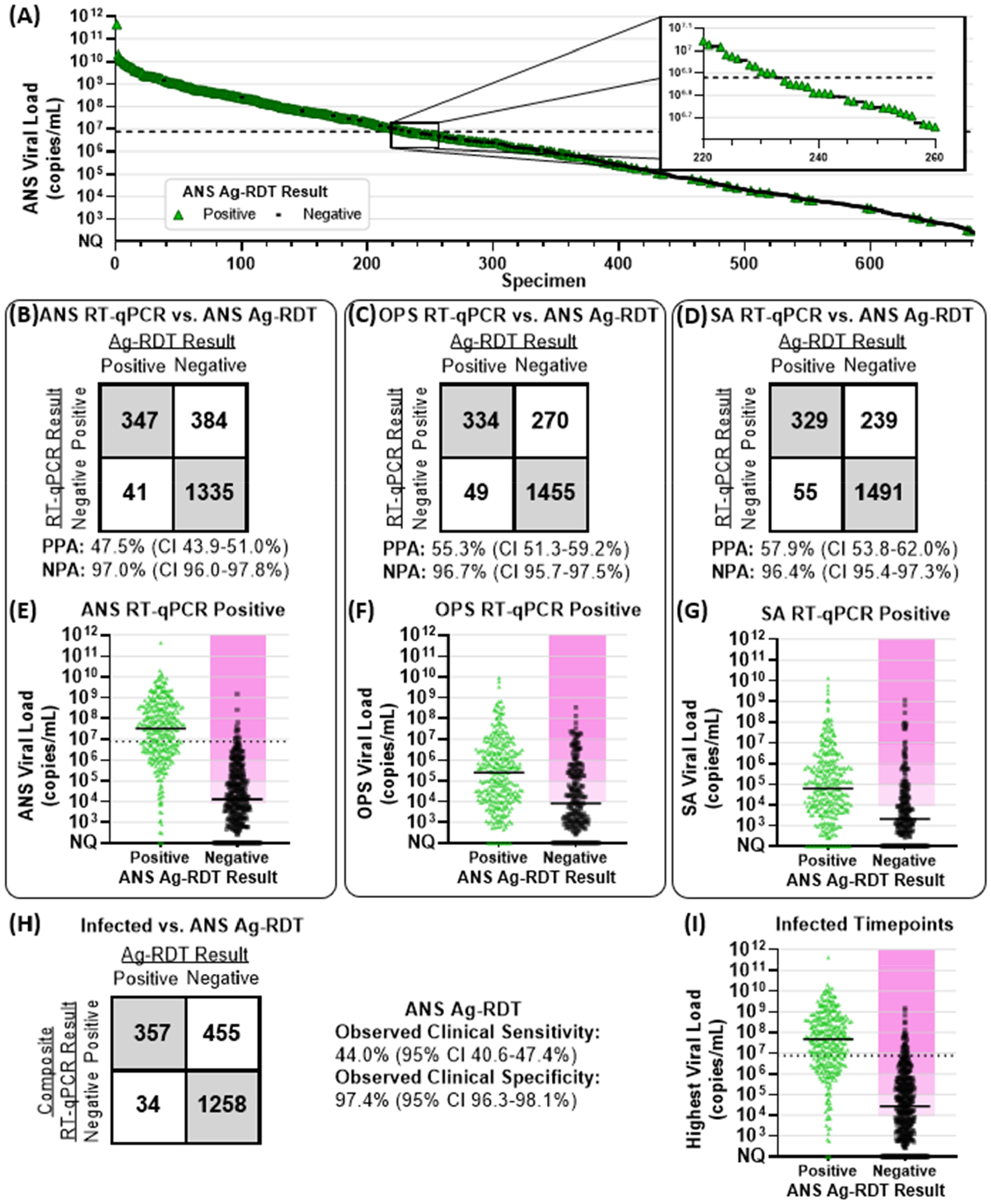
Comparison of Anterior-Nares Swab Antigen Rapid Diagnostic Test (Ag-RDT) Results to RT-qPCR Results and Viral Loads. **(A)** 680 ANS specimens with quantifiable SARS-CoV-2 viral loads are ordered by viral load and colored by Ag-RDT results (green for positive antigen test result, black for antigen negative). Inset shows higher resolution for results with viral loads around 7.6×10^6^ copies/mL (black dashed line), above which 95% of ANS specimen resulted Ag-RDT positive. 2×2 matrices of concordance between ANS Ag-RDT results and valid, conclusive RT-qPCR results for **(B)** 2107 ANS specimens, **(C)** 2108 OPS specimens and **(D)** 2114 SA specimens. PPA, positive percent agreement; NPA, negative percent agreement. CI indicates 95% confidence interval. Distribution of viral loads from **(E)** 731 RT-qPCR positive ANS specimens, **(F)** 604 RT-qPCR positive OPS specimens and **(G)** 568 RT-qPCR positive SA specimens, with either positive or negative Ag-RDT results. Solid horizontal black lines indicate medians. **(H)** A 2×2 matrix of observed concordance between Ag-RDT results, and infected status, based on composite RT-qPCR results from all three specimen types, at 2104 timepoints with valid, conclusive results for all specimen types by RT-qPCR and valid ANS Ag-RDT results. **(I)** Distribution of the highest viral load among ANS, OPS, and SA specimens collected by any participant at 812 composite RT-qPCR positive (infected) timepoints, with either positive or negative Ag-RDT results. Magenta shading in panels E, F, G, and I indicates infectious viral loads (above 10^4^, 10^5^, 10^6^ or 10^7^ copies/mL). ANS, anterior-nares swab; OPS, oropharyngeal swab; SA, saliva; Ag-RDT, antigen rapid diagnostic test. Detailed tabulation including inconclusive and invalid results shown in **Table S2**.

**Figure 3.**
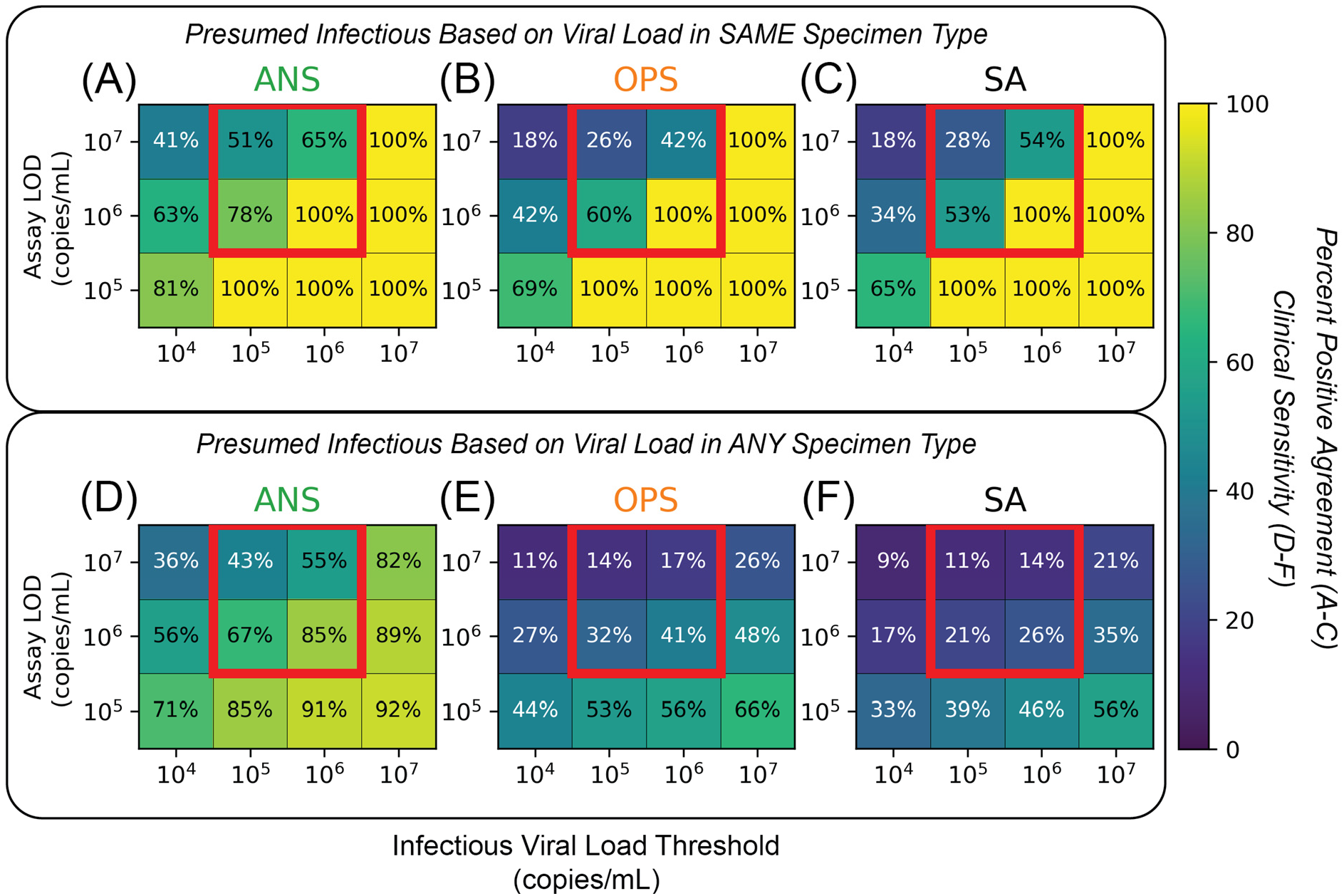
Effects of Low-Analytical-Sensitivity Assay LOD, Infectious Viral-Load Threshold (IVLT), and Inclusion of Multiple Specimen Types on Inferred Clinical Sensitivity to Detect Presumed Infectious Individuals. **(A-C)** Heatmaps visualizing positive percent agreement (A-C) for each specimen type, **(A)** saliva (SA), **(B)** anterior-nares swab (ANS), and **(C)** oropharyngeal swab (OPS), tested with assays of different LODs in the range of low-analytical-sensitivity tests (such as Ag-RDTs) to detect individuals presumed infectious only if the viral load in the tested specimen type was at or above a given IVLT. Red boxes highlight an important interaction between assay LOD and IVLT that is elaborated in the text. **(D-F)** Heatmaps visualizing the inferred clinical sensitivity for each specimen type, **(D)** SA, **(E)** ANS, **(F)** OPS, tested with assays of different LODs to detect individuals presumed to be infectious if the viral load in any specimen type was at or above a given IVLT. Heatmaps for computationally contrived combination specimen types are shown in **Figure S5**.

**Figure 4.**
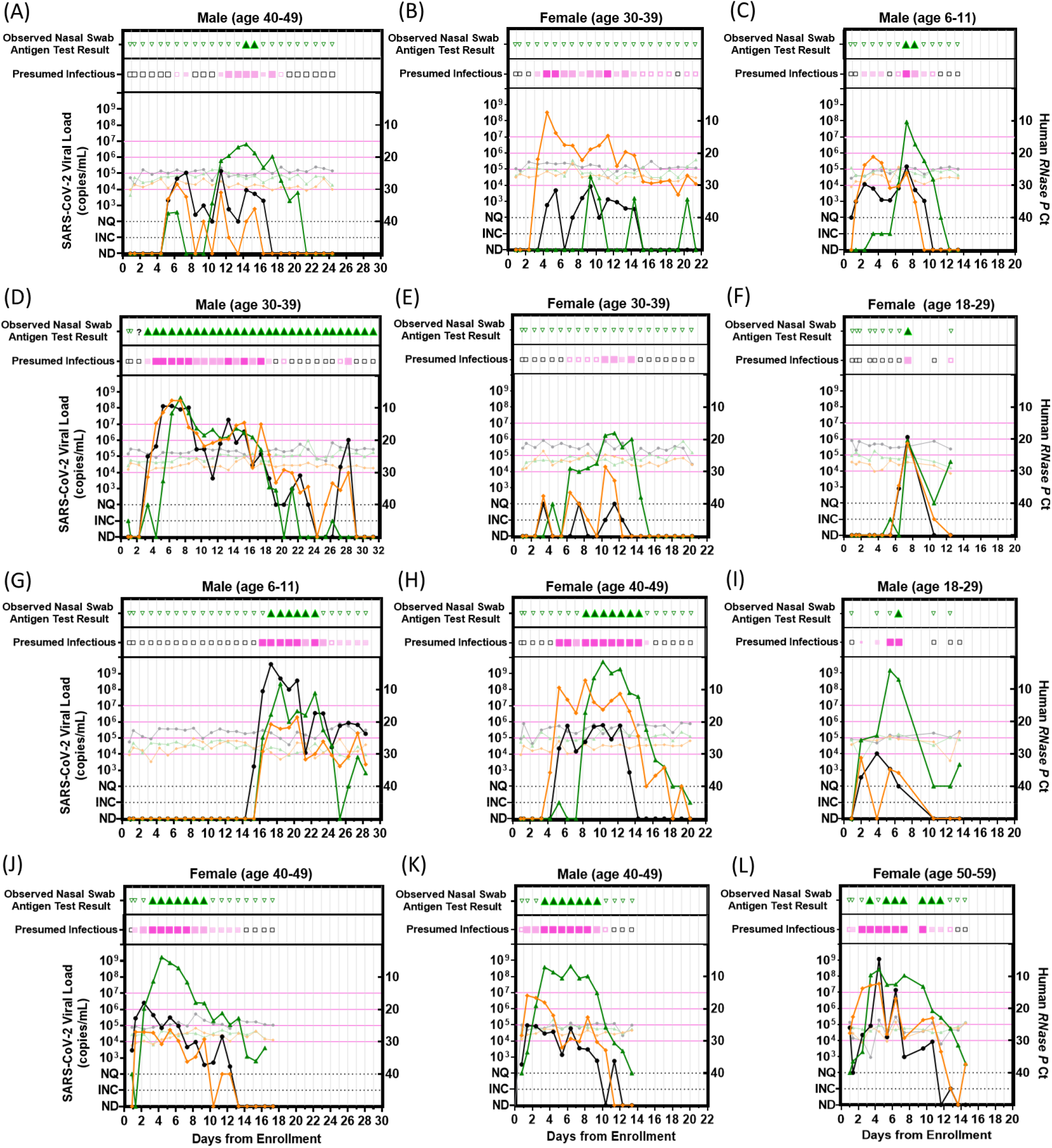
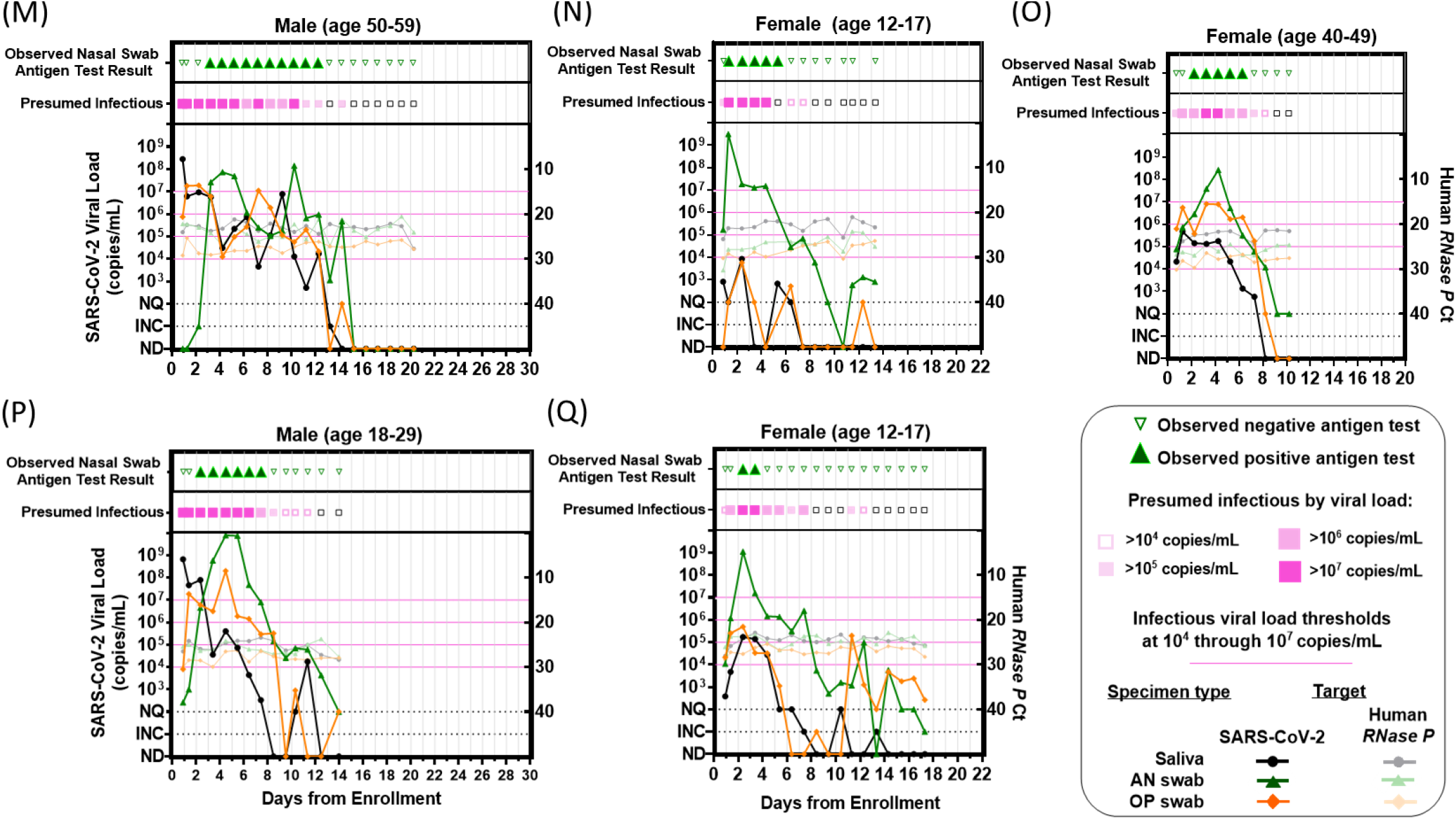
Longitudinal Viral Loads and Antigen Rapid Diagnostic Testing. Each panel **(A-Q)** represents a single participant throughout the course of enrollment, with observed ANS rapid antigen testing results, presumed infectious period (magenta) based on viral loads at or above each infectious viral-load threshold 10^4^ to 10^7^ copies/mL in any specimen type, SARS-CoV-2 viral loads (left y-axis) and human *RNase P* Ct values (right y-axis) by RT-qPCR in each specimen type. Viral-load data for participants A-N were reported previously.^55^ INC, inconclusive; NQ, viral load detected but below the test LOD (250 copies/mL); ND, not detected for RT-qPCR measurements; AN, anterior-nares; OP, oropharyngeal. A single invalid antigen test is indicated with a “?” symbol in panel D.

**Figure 5.**
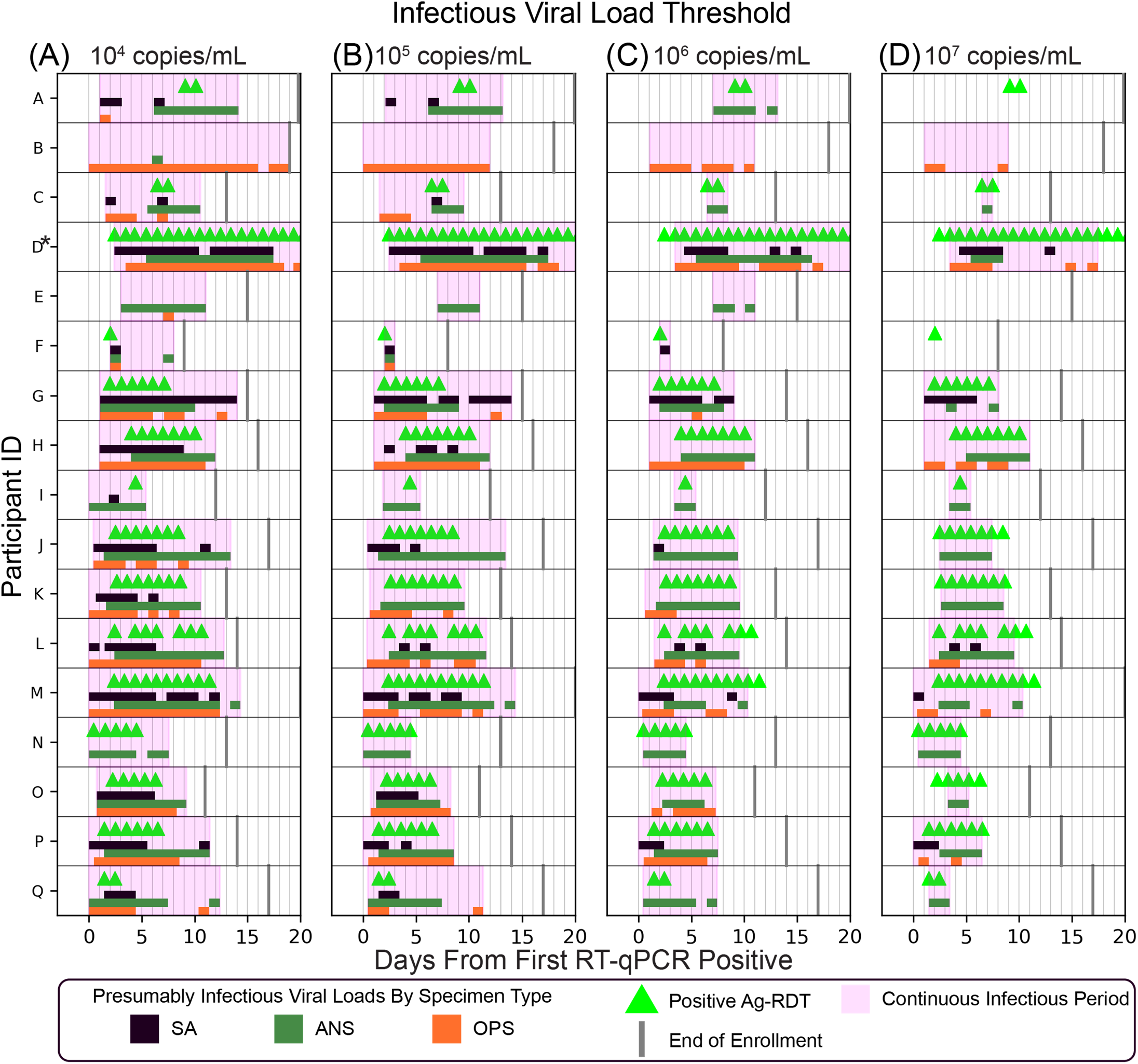
Periods of Presumed Infectiousness as a Factor of Infectious Viral-Load Threshold (IVLT). **(A-D)** Days starting from first RT-qPCR positive that each participant (A-Q; see **Fig 3**) had presumably infectious viral loads (with IVLTs of 10^4^ to 10^7^ copies/mL) in each specimen type (green bars, anterior nares swab [ANS]; orange bars, oropharyngeal swab [OPS]; black bars, saliva [SA]). Positive Ag-RDT tests are indicated with green triangles and the final date of study enrollment for each is indicated with grey lines. The timecourse for participant D (who experienced a series of false-positive antigen tests) is truncated, indicated by a * (see Supplementary Information).

**Figure 6.**
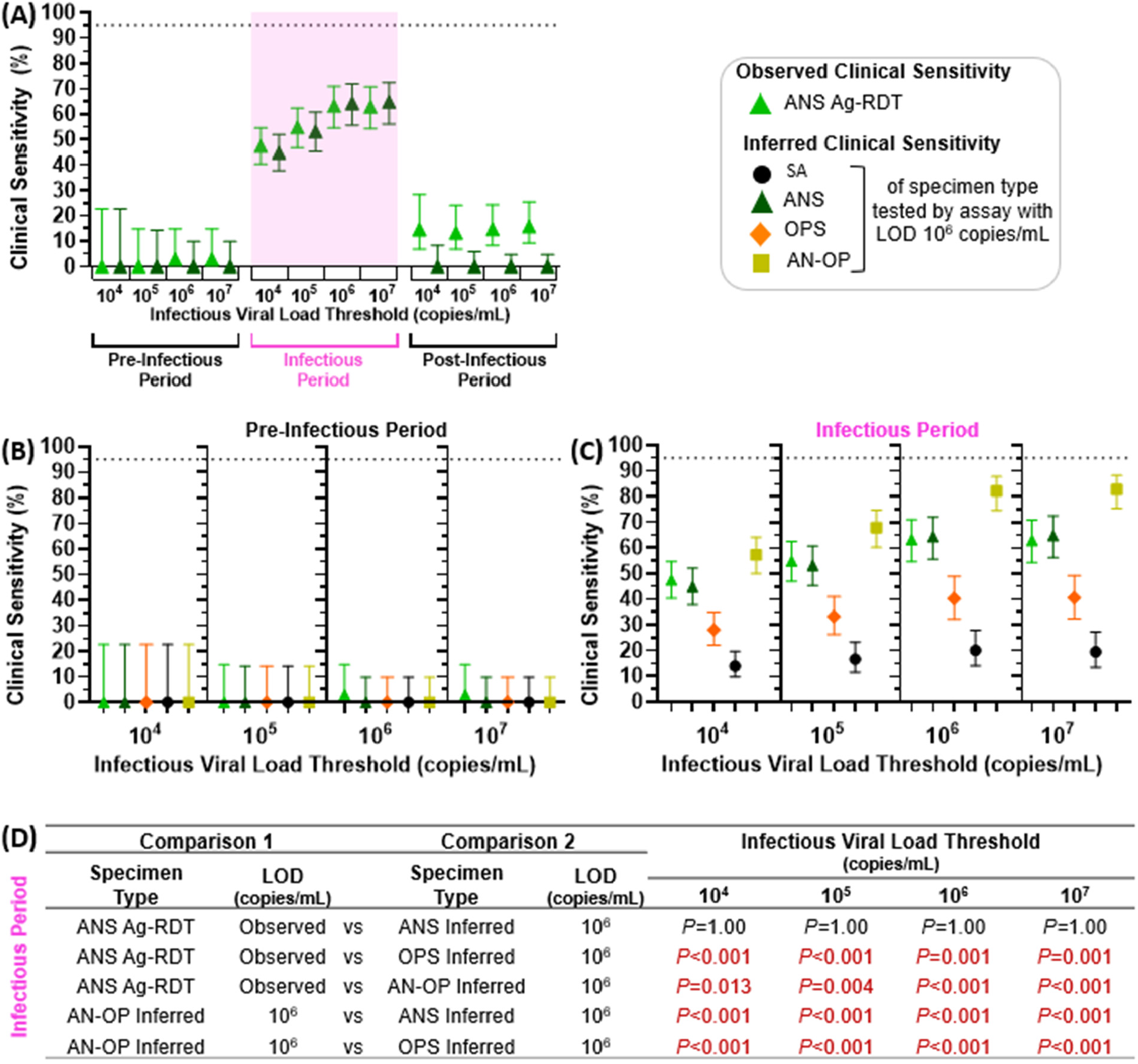
Observed and Inferred Performance of Low-Analytical-Sensitivity Daily Antigen Rapid Diagnostic Tests (Ag-RDTs) to Detect Presumed Infectious Individuals. Individuals were presumed infectious for the period between first specimen (of any type) with a viral load above the IVLT (10^4^, 10^5^, 10^6^, or 10^7^ copies/mL) until all specimen types were below the IVLT; specimens collected prior to this period were considered pre-infectious, and after this period, post-infectious. **(A)** Observed clinical sensitivity of the ANS Ag-RDT (fluorescent green), and the inferred clinical sensitivity of an ANS test with an LOD of 10^6^ copies/mL (green), for each stage of infection. Subsequent plots show the observed clinical sensitivity for detection of presumed infectious individuals by the ANS Ag-RDT (fluorescent green) and the inferred clinical sensitivity for ANS (green), OPS (orange), SA (black), and a computationally-contrived AN–OP combination swab specimen type (yellow) during the **(B)** pre-infectious period and **(C)** infectious period of infection. Inferred clinical sensitivity was based on measured viral loads in the given specimen type at or above an LOD of 10^6^. **(D)** Comparison of clinical sensitivities using McNemar Exact Test, for given comparisons across specimen type. ANS Ag-RDT vs ANS with LOD 10^6^ copies/mL was tested using a two-tailed McNemar Exact Test; all other combinations use a one-tailed McNemar exact test. *P*-values were adjusted using a Benjamini–Yekutieli correction to account for multiple hypotheses being tested. Additional analyses of the inferred clinical sensitivity of the anterior-nares—oropharyngeal combination swab can be found in a separate manuscript.^55^ SA, saliva; ANS, anterior-nares swab; OPS, oropharyngeal swab; AN–OP, anterior-nares–oropharyngeal combination swab; LOD, limit of detection.

Viral sequencing for variant determination was performed on an ANS or OPS specimen with high viral load from at least one infected individual in each household, as previously described.^55^ **Fig 7** was created using BioRender.

**Figure 7.**
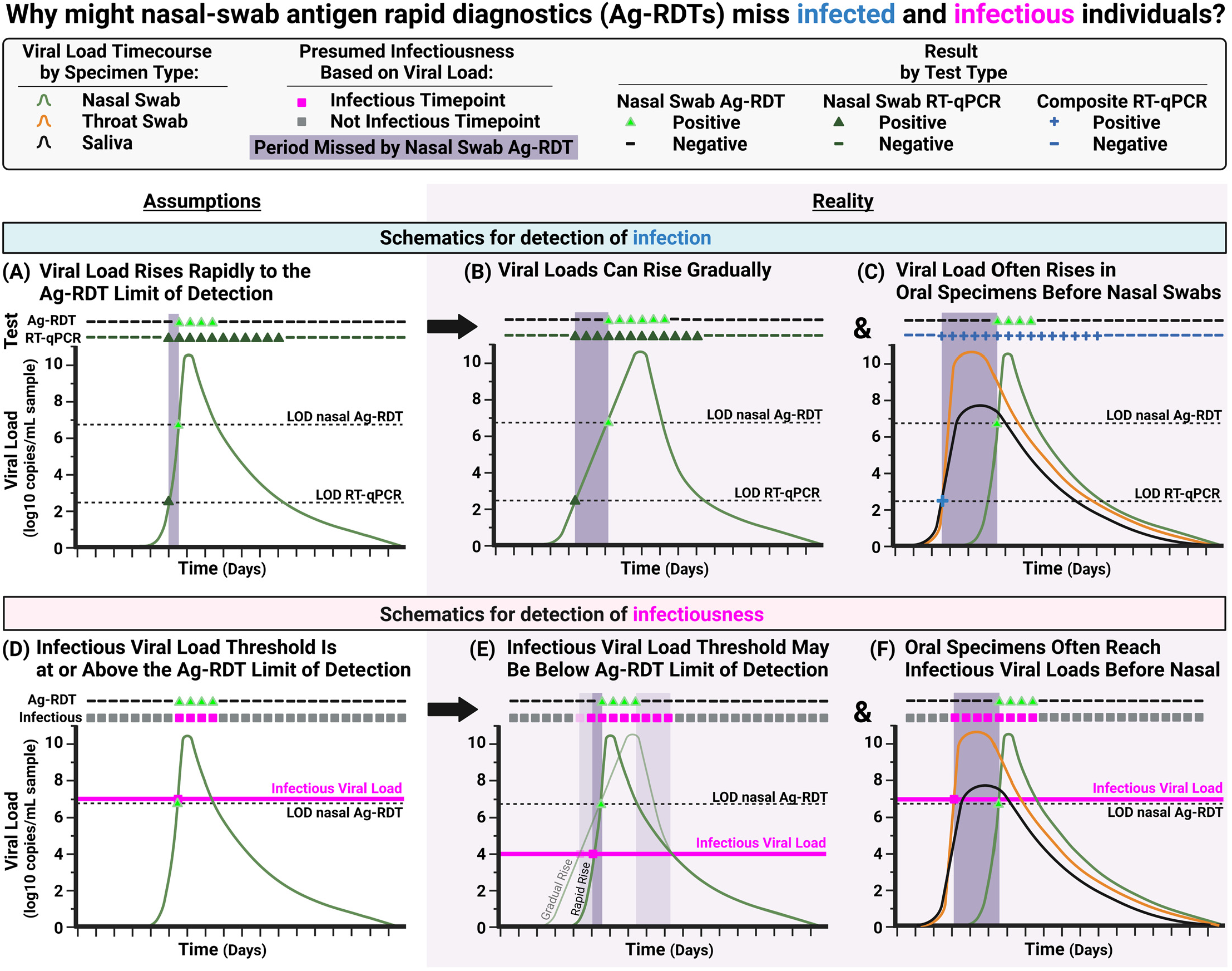
Conceptual Diagrams Illustrating Why Nasal-only Antigen Rapid Diagnostic Tests (Ag-RDTs) Are Likely to Miss Infected and Infectious Individuals. **(A)** Schematic of an idealized, hypothetical viral-load timecourse in which viral load rises quickly from detectable to the limit of detection (LOD) of a low-analytical-sensitivity test, such as Ag-RDTs. Such a pattern would result in a daily Ag-RDT being effective for detection of infection (diagram based on a commonly held view^27^). **(B)** Schematic of a viral-load timecourse based on longitudinal viral-load data^31,41,43,47-58^ in which, for some individuals, early viral loads rise gradually, resulting in detection of the infected individual several days earlier by a high-analytical-sensitivity test than by the Ag-RDT. This mechanism for missed detection by COVID-19 Ag-RDTs has been previously hypothesized.^59^ **(C)** Schematic of a viral-load timecourse based on observed paired, longitudinal viral-load data in which individuals exhibit a rise in viral load in oral (saliva or throat swab) specimens days before viral loads rise in nasal specimens.^41,43,49,51,52,55^ When these additional specimen types are used to assign a composite infection status, the nasal Ag-RDT is revealed to have poor performance. **(D)** Schematic of an idealized, hypothetical viral-load timecourse (based on commonly held views^27,48,81^) in which viral load rises quickly from detectable to infectious, and the infectious viral load threshold is equivalent to the LOD of the Ag-RDT. Such a pattern would result in near-perfect detection of infectious individuals by the daily Ag-RDT. **(E)** Schematic of a viral-load timecourse in which the infectious viral load is lower than the LOD of the Ag-RDT. Here, infectious individuals would be missed by the Ag-RDT during the period viral load is between the infectious viral load threshold and the LOD of the Ag-RDT. This period will be longer if the rise in viral load is gradual (light-green line) rather than quick (dark-green line). **(F)** Schematic of a viral-load timecourse in which individuals exhibit high, presumably infectious viral loads in saliva or throat swab specimens while nasal swab viral loads remain very low, particularly at the beginning of infection. Here, the ANS Ag-RDT is unable to detect most infectious timepoints. The dashed LOD nasal Ag-RDT line indicates the inferred LOD for the nasal-swab Ag-RDT we evaluated (7.6 × 10^6^ copies/mL). The dashed LOD RT-qPCR line indicates the LOD of the RT-qPCR assay used in this study (250 copies/mL). The pink Infectious Viral Load line indicates a threshold associated with the presence of replication-competent virus; individuals are considered infectious if any specimen type has a viral load above the threshold.

### Statistical Analyses

Positive and negative percent agreement for each specimen type (**Fig 2A-C, Fig 3A-C**) was calculated as the number of specimens with concordant results by RT-qPCR and ANS Ag-RDT over the total number of specimens with positive or negative results, respectively, by RT-qPCR for the given specimen type as reference test.

We used quantitative viral loads to predict whether an assay with a given analytical sensitivity would result positive or negative. If the viral load in the specimen type was above a given limit of detection (LOD) for a hypothetical assay, the result was predicted to be positive, whereas if viral loads were below the LOD for a hypothetical assay, the result was predicted to be negative. We also predicted results of a computationally-contrived AN-OP combination swab specimen type (**Fig 6**), using the higher viral load of either the ANS or OPS specimens from the participant at a given timepoint, as previously described.^55^ Predicted results were used to calculate inferred positive percent agreement (**Fig 3A-C)** or inferred clinical sensitivity **(Fig 5, Fig 6**).

Clinical sensitivity (**Fig 2H, Fig 3D-F, Fig 5, Fig 6**) was calculated as the number of specimens with either observed or predicted positive results (based on viral loads above a specified assay LOD) over the total number of infected or infectious timepoints included. We denote clinical sensitivity as inferred when predicted based on quantitative viral loads.

We also used quantitative viral-load measurements to presume whether an individual was infectious at a given timepoint (**Fig 3, Fig 4, Fig 5, Fig 6**). Individuals were presumed to be infectious at all timepoints between the first timepoint where at least one specimen type had a viral load above a specified infectious viral-load threshold (IVLT) of 10^4^, 10^5^, 10^6^, or 10^7^ copies/mL (based on viral culture literature^16,25,30-47^), until the last timepoint where at least one specimen type had a viral load above the IVLT.

Confidence intervals were calculated as described in the Clinical Laboratory Standards Institute EP12-A2 User Protocol for Evaluation of Qualitative Test Performance.^78^ RT-qPCR results and ANS Ag-RDT results were considered paired. Differences in the inferred or observed clinical sensitivity from these paired data were tested for statistical significance (**Fig 6I**) using the McNemar exact test performed using the statsmodels package in Python v3.8.8, with a Benjamini–Yekutieli procedure to correct *P*-values.

## RESULTS

### Ag-RDT Exhibits <50% Observed Clinical Sensitivity to Detect Infected Individuals

We first performed a cross-sectional analysis to estimate the limit of detection (LOD) of the ANS Ag-RDT, then compare the positive percent agreement (PPA) of the ANS Ag-RDT against ANS RT-qPCR or composite infection status based on RT-qPCR results from ANS, OPS and SA. Of 680 ANS specimens with quantifiable viral loads and valid, paired ANS Ag-RDT results, 95% positive percent agreement (PPA) was observed when ANS specimens had viral loads above 7.6×10^6^ copies/mL (**Fig 2A**), suggesting this value as an estimate for the approximate, inferred LOD of the assay. The approximately 1000-fold difference between RNA viral load and viral titer reported by the manufacturer^74^ is reasonably expected.^16,48^ We observed 48% (347 of 731) PPA between the ANS Ag-RDT and ANS RT-qPCR (**Fig 2B**). However, the observed clinical sensitivity of the ANS Ag-RDT (**Fig 2H**) when compared to composite infection status (based on RT-qPCR results from any of the three specimen types tested) was 44% (357 of 812 infected timepoints), significantly lower (*P*<0.001, upper-tailed McNemar exact test) than the PPA of the ANS Ag-RDT against ANS RT-qPCR alone (**Fig 2B**). Although a low PPA and clinical sensitivity are expected due to the difference in analytical sensitivity between the Ag-RDT and RT-qPCR, many timepoints during which participants had high, presumably infectious viral loads in either ANS, SA, or OPS specimens resulted negative by the ANS Ag-RDT (**Fig 2E-G**). Approximately 50% of timepoints at which the ANS Ag-RDT resulted negative had viral loads above 10^4^ copies/mL in ANS (**Fig 2E**) or OPS (**Fig 2F**) or any specimen type (**Fig 2I**).

### Assay Analytical Sensitivity, IVLT, and Consideration of Multiple Specimen Types Strongly Impacts Clinical Sensitivity to Detect Infectious Individuals

We next sought to assess how well presumably infectious individuals were detected by the Ag-RDT. Because high viral loads can be used to classify individuals as likely infectious, we first examined how the selection of an infectious viral load threshold (IVLT) impacts the inferred clinical sensitivity of low-analytical-sensitivity assays (such as Ag-RDTs) to detect presumed infectious individuals. We created a matrix of IVLTs (10^4^, 10^5^, 10^6^ or 10^7^ copies/mL) and low-analytical-sensitivity assay LODs (10^5^, 10^6^, or 10^7^ copies/mL), for each specimen type. In each cell, we calculated the inferred clinical sensitivity of the hypothetical assay to detect timepoints with viral loads above the IVLT. We calculated the inferred clinical sensitivity against timepoints with viral loads above the IVLT only in that specimen type (**Fig 3A-C**), and against timepoints with viral loads above the IVLT in any of the three specimen types (**Fig 3D-F**).

When considering viral loads only in the specimen type tested, clinical sensitivity increases as the IVLT increases, and decreases as assay LOD increases. Setting an IVLT at or above the LOD of the assay artificially increased the inferred clinical sensitivity to detect presumed infectious individuals. We highlight three instances (red boxes in **Fig 3A-C**) where inferred clinical sensitivities increased by up to 74% as a result of changing the IVLT. Perfect performance was observed in the lower-right cells of the heatmap (**Fig 3A-C**) where the IVLT was at or above the assay LOD. This analysis demonstrated how selection of an IVLT similar to the assay LOD can cause gross overestimates of clinical sensitivity to detect infectious individuals.

Importantly, when considering viral loads above the IVLT in any of the three specimen types tested (**Fig 3D-F**), inferred clinical sensitivities decreased for all specimen types, regardless of IVLT or assay LOD. Because of extreme differences in viral load that can exist among specimen types from the same individual at a given timepoint,^55^ individuals often have high, presumably infectious viral loads in one but not all specimen types. For that reason, inferred clinical sensitivity decreases drastically when the potential for infectiousness in multiple specimen types, rather than just one, is considered.

### Longitudinal Ag-RDT Performance

Cross-sectional analyses (**Fig 2, Fig 3**) cannot assess performance of the Ag-RDT at different periods of acute infection, including early detection. Thus, we next assessed the performance of the ANS Ag-RDT longitudinally throughout the course of the infection. We identified a cohort of 17 individuals who began sampling very early in the course of acute infection (**Fig 1**) from whom we could assess Ag-RDT performance through different periods of SARS-CoV-2 infection. We compiled each participant’s daily viral-load measurements (and human *RNase P* Ct values to assess sampling consistency) for each specimen type (SA, ANS, OPS),^55^ as well as their paired daily ANS Ag-RDT results, and classified whether the participant was presumably infectious at each timepoint based on whether viral loads in any of their three specimen types were above a given IVLT (**Fig 4**).

All but two of the 17 participants (**Fig 4D**,**F**) reached presumed infectious viral loads at least 1 day prior to their first positive ANS Ag-RDT result: Of these 15 participants, 6 had a delay of 1-2 days (**Fig 4G**,**J**,**N**,**O**,**P**,**Q**), 5 had a delay of 3 days (**Fig 4H**,**I**,**K**,**L**,**M**), 1 had a delay of 5 days (**Fig 4C**) and 1 had a delay of 8 days (**Fig 4A**). Two participants (**Fig 4B**,**E**) had infectious viral loads for more than 8 days each, but neither ever reported a positive ANS Ag-RDT result, one of whom (**Fig 4B**) had high (>10^5^ copies/mL) viral loads in OPS specimens for 12 days while ANS specimens remained at low levels (rising just above 10^4^ copies/mL only once). The other participant (**Fig 4E**) had ANS viral loads >10^6^ copies/mL on 3 days, but never yielded a positive Ag-RDT result, likely because these viral loads were just below the ANS Ag-RDT LOD. In this cohort, the overall observed clinical sensitivity of the ANS Ag-RDT to detect infected individuals was significantly higher when participants were symptomatic than when asymptomatic (**Fig S1**), but low (<50%) at both symptomatic and asymptomatic timepoints.

### Nasal Swab Ag-RDT Misses Infectious Viral Loads That Occur Asynchronously in Saliva and Oropharyngeal Swabs

Given that many individuals exhibited a delay between high, presumably infectious viral loads and their first ANS Ag-RDT positive result (**Fig 4**), we wanted to assess how the periods of infectious viral loads in each of the three specimen types overlapped with each other, and which infected and infectious timepoints were detected by the ANS Ag-RDT.

First, we visualized the timing of the infectious periods based on viral loads (above IVLTs of 10^4^, 10^5^, 10^6^ or 10^7^ copies/mL) in each specimen type, alongside ANS Ag-RDT results. Each participant’s infection was aligned to the first RT-qPCR positive result in any specimen type. Then, for each IVLT, we plotted the periods when each specimen type had viral loads above the IVLT. The period of viral loads above the IVLT in any specimen type was indicated in magenta, and positive ANS Ag-RDT results were overlaid (**Fig 5**).

For IVLTs below 10^7^ copies/mL (**Fig 5A-C**), all 17 individuals were presumed to be infectious for at least one day of the infection. As IVLT was increased, the length of the infectious period for each participant decreased. At an IVLT of 10^7^ copies/mL (**Fig 5D**), three participants (**Fig 5D[A**,**E**,**F])** would not be considered infectious.

If the infectious periods in OPS and SA overlapped perfectly with the infectious period in ANS, then infectious viral loads in OPS or SA would not affect the performance of the ANS Ag-RDT to detect infectious individuals. However, we find this is not the case. The presumed infectious periods for different specimen types were often asynchronous (non-overlapping): for many individuals OPS and SA specimens reached infectious viral loads prior to infectious viral loads in ANS. For this reason, the ANS Ag-RDT often resulted negative during the infectious period (pink-shaded days lacking green triangles in **Fig 5**), particularly during the early part of the infectious period.

### Performance of Daily Nasal Swab Ag-RDT to Detect the Pre-Infectious and Infectious Periods

We next investigated the performance of the daily ANS Ag-RDT to detect individuals during the pre-infectious and infectious periods. We separately analyzed IVLTs of 10^4^ to 10^7^ copies/mL and plotted the observed clinical sensitivity of the ANS Ag-RDT alongside the inferred clinical sensitivity of ANS specimens when tested by an assay with a similar LOD (10^6^ copies/mL).

The inferred clinical sensitivity predicted for ANS specimens tested by a hypothetical assay with LOD of 10^6^ copies/mL and the observed clinical sensitivity of the Ag-RDT was similar for both the pre-infectious and infectious periods, and across all four IVLTs (**Fig 6A**,**D**). This congruency supported the use of quantitative viral loads to predict Ag-RDT performance.

In the pre-infectious period, the ANS Ag-RDT was positive in, at most, 1 of 34 timepoints (**Fig 6B**). Increasing the IVLT decreased the length of the infectious period, which increased the observed clinical sensitivity of the ANS Ag-RDT to detect infectious individuals (**Fig 6C**). However, even at the highest IVLT (10^7^ copies/mL), the ANS Ag-RDT detected only 63% of presumed infectious individuals (**Fig 6C**).

We also inferred the clinical sensitivity of other specimen types (SA, OPS, and a computationally-contrived combination AN-OP swab specimen type) if tested by an assay with similar analytical sensitivity as the ANS Ag-RDT. We inferred that if tested by an assay with LOD of 10^6^ copies/mL, no single specimen type (ANS, OPS, SA) would achieve 95% clinical sensitivity to detect infectious individuals, for any IVLT (**Fig 6C**). However, a computationally-contrived AN–OP combination swab specimen at an LOD of 10^6^ copies/mL was predicted to perform significantly better than any single specimen type, including the observed performance of the ANS Ag-RDT we evaluated (**Fig 6D**). At this low analytical sensitivity, though, the combination AN-OP combination swab was unable to detect individuals in the pre-infectious period of infection.

## CONCLUSIONS

Our results revealed three key findings relevant to the use of Ag-RDTs and, by extension, other tests with low and moderate analytical sensitivity (including some molecular tests that forgo nucleic acid extraction and purification).

First, our field evaluation of an ANS Ag-RDT showed low (44%) clinical sensitivity for detecting infected persons at any stage of infection. This clinical sensitivity is consistent with another recent field evaluation of this ANS Ag-RDT when used for twice-weekly screening testing on a college campus.^23^ It is also consistent with FDA^79^ and CDC guidance^80^ that using two or more repeat ANS Ag-RDTs are needed to improve the clinical sensitivity of these tests.

There are two reasons for the observed low clinical sensitivity of the ANS Ag-RDT to detect infected individuals: (i) First, the low analytical sensitivity of Ag-RDTs requires high viral loads to yield a positive result. Although it has been proposed^27^ that a rapid rise in viral load reduces the advantage of tests that can detect low viral loads (**Fig 7A**), this advantage remains when there is a more gradual rise in viral loads (**Fig 7B**) as we observed in some individuals (**Fig4A,C**). (ii) The second, and more impactful reason, is that many early-infection timepoints had detectable virus in saliva and throat swabs, but not nasal swabs. A nasal-swab reference test would miss these infected timepoints. Therefore, the true performance of an ANS Ag-RDT would be worse when compared to composite infection status based on multiple upper-respiratory specimen types than nasal-swab alone (**Fig 7C**).

These two reasons for poor detection of infected individuals by Ag-RDTs have important implications for the design and interpretation of other Ag-RDT evaluations. Because viral-load timecourses in different specimen types from an individual are asynchronous,^55^ the true clinical sensitivity of an Ag-RDT to detect infected individuals will be lower than reported in diagnostic-test field evaluations that compare only to an ANS reference test.^22,50,66,69,70,72,73,82 21^ Relatedly, the positive percent agreement reported by the manufacturer to the FDA for authorization of this Ag-RDT (83.5%) was calculated relative to detection by a RT-PCR reference test on only nasal swabs—nearly all of which (84 of 91) were collected from individuals after symptom onset.^74^ Our work suggests that governing bodies, such as the FDA, should require that trials evaluating the clinical sensitivity of an Ag-RDT to detect infected individuals based on a composite infection status from multiple anatomical upper-respiratory specimen types.

Our second key finding is that the ANS Ag-RDT also poorly detected individuals presumed to be infectious based on high viral loads. The ANS Ag-RDT detected only up to 3% of pre-infectious and only up to 63% of presumed infectious timepoints. This low clinical sensitivity to detect infectious individuals was not consistent with a common view^27^ that proposes low-analytical-sensitivity tests will have near-perfect detection of infectious individuals (**Fig 7D**). Our data demonstrate that this common but idealized view misses two important points. (i) First, in the common view, the LOD of the Ag-RDT aligns with the infectious viral load threshold (IVLT) (**Fig 7D**), though there is no fundamental reason why the LOD should align perfectly with the IVLT. Replication-competent virus has been reliably isolated from specimens with viral loads of 10^4^ copies/mL and above,^25,32-40^ whereas the LOD of Ag-RDTs span several orders of magnitude (from approximately 10^5^ copies/mL to 10^7^ copies/mL). The Ag-RDT we evaluated in this study was inferred to have an approximate LOD of 7.6×10^6^ copies/mL. As our data demonstrate (**Fig 3**), if the chosen IVLT is at or above the LOD of the test, the test will be predicted to have near perfect clinical sensitivity to detect presumed infectious individuals. However, if the true IVLT is below the LOD, calculated clinical sensitivity may be reduced by up to 56%. Therefore, a difference between the true IVLT and the LOD of the test would result in lower performance of the Ag-RDT to detect infectious individuals (**Fig 7E**). Additionally, if viral loads rise gradually, there will be more time between when an individual becomes infectious and when viral loads become detectable by the Ag-RDT. (ii) The second point that the common, idealized view misses is the potential for asynchronous infectious virus in specimen types other than the one tested by the Ag-RDT. We observed infectious viral loads in saliva and throat swab specimens at all IVLTs from 10^4^ to 10^7^ copies/mL, even while nasal swab viral loads were very low, well below the LOD of Ag-RDT. As expected, the ANS Ag-RDT was unable to detect presumably infectious individuals at these timepoints. For example, in one individual (**Fig 4A**), nasal-swab viral loads were absent or below 10^3^ copies/mL for the first 5 days of infection, resulting in negative ANS Ag-RDT results despite high and presumably infectious viral loads in multiple SA and OPS specimens from this individual. Because the ANS Ag-RDT can only detect individuals with high, presumably infectious viral loads in nasal swabs, individuals with infectious virus in other specimen types will be missed (**Fig 7F**).

Missing these two important points has several critical implications for evaluating an Ag-RDT’s ability to detect infectious individuals. Some agent-based outbreak models^,9,25,83,84^have inferred that low-analytical-sensitivity tests would be effective at mitigating SARS-CoV-2 transmission in a population. Individuals (or agents) in these models^9,25,83-87^ are considered infectious and capable of transmitting to other agents when viral loads are above a chosen IVLT. These models may overestimate test effectiveness if infectiousness is based only on simulated viral loads in a single, tested specimen type, and/or if the IVLT chosen is similar to the LOD of the simulated test. Additionally, nearly all studies evaluating Ag-RDT concordance with infectiousness performed viral culture only on a single specimen type^20,21,41,46,48-50,63,65-70,88^ potentially overlooking infectious virus in other specimen types. One of these studies^65^ is cited as basis for CDC^80^ recommendations to use of repeat ANS Ag-RDTs to improve the clinical sensitivity of these tests.

Our third key finding is that the use of combination specimen types can significantly improve the performance of Ag-RDTs. Our data suggest that using an AN–OP combination swab specimen type tested by an assay with a similar LOD as the ANS Ag-RDT we directly evaluated would significantly improve detection of infected and infectious individuals. Improved detection with an AN–OP combination swab for a different Ag-RDT was recently demonstrated for asymptomatic individuals at a testing center^89^ and has been used to evaluate a panel of Ag-RDTs.^34^ Many countries have already authorized and/or implemented the use of combination specimen types, including for Ag-RDTs,^4,90,91^ yet this is not the case in the U.S., where all at-home Ag-RDTs use nasal swabs.

We acknowledge several limitations of our study. First, we only evaluated a single Ag-RDT, the Quidel QuickVue At-Home OTC COVID-19 Test. Although other Ag-RDTs have different LODs,^10,76,77^ equivalence between the clinical sensitivity of this Ag-RDT directly observed versus inferred based on nasal swab viral loads supports that performance of other Ag-RDTs can be inferred from quantitative viral-load data. Second, we infer the clinical sensitivity of a combination AN–OP swab based on viral-load measurements, but did not directly test a combination swab in this study population. Finally, this study was performed in the context of two SARS-CoV-2 variants (Delta and Omicron) and in one geographical area. As new SARS-CoV-2 variants emerge that may demonstrate different viral-load kinetics, it will be critical to re-evaluate testing strategies to effectively detect pre-infectious and infectious individuals.

Ag-RDTs are useful tools for rapid identification of individuals with high viral loads in the specimen type tested. As discussed above, the utility of Ag-RDTs for detection of infected and presumably infectious individuals is often justified using several assumptions (**Fig 7**), in particular using the assumption that at a given timepoint the viral loads in all specimen types from an individual are similar. Our study demonstrates that this assumption is not justified. Re-evaluating these assumptions based on new evidence can inform more effective testing strategies, both for SARS-CoV-2 and for other respiratory viral pathogens.

## Data Availability

The data underlying the results presented in the study can be accessed via CaltechDATA: https://data.caltech.edu/records/20223.

## Acknowledgments

We sincerely thank the study participants for making this work possible. We thank Lauriane Quenee, Grace Fisher-Adams, Junie Hildebrandt, Megan Hayashi, RuthAnne Bevier, Chantal D’Apuzzo, Ralph Adolphs, Victor Rivera, Steve Chapman, Gary Waters, Leonard Edwards, Gaylene Ursua, Cynthia Ramos, and Shannon Yamashita for their assistance and advice on study implementation and/or administration. We thank Jessica Leong, Ojas Pradhan, Si Hyung Jin, Emily Savela, Bridget Yang, Ekta Patel, Hsiuchen Chen, Paresh Samantaray, Zeynep Turan, Cindy Kim, Trinity Lee, Vanessa Mechan, Katherine Stiefel, Rosie Zedan, Rahulijeet Chadha, Minkyo Lee, and Jenny Ji for volunteering their time to help with this study. We thank Prabhu Gounder, Tony Chang, Jennifer Howes, and Nari Shin for their support with recruitment. Finally, we thank all the case investigators and contact tracers at the Pasadena Public Health Department and Caltech Student Wellness Services for their efforts in study recruitment and their work in the pandemic response.

## Author contributions

*Detailed author contributions are in the Supplement*.

Conceptualization: MF, Y-YG, RFI, NS, AVW

Methodology: RA, NS, AVW

Investigation: RA, AMC, YCC, SC, HD, MKK, JRBR, AER, NS, AVW, TY

Visualization: RA, NS, AVW

Funding acquisition: RFI, AVW

Project administration: RFI, NS

Supervision: YCC, RFI

Writing – original draft: RA, NS, AVW

Writing – review & editing: RA, AMC, RFI, AER, NS, AVW

## Data and materials availability

The data underlying the results presented in the study can be accessed at CaltechDATA: https://data.caltech.edu/records/20223.

## Supplementary Information

This article contains a supplementary information PDF.

## Supplemental Information for

### Supplementary Methods

#### Study Population

This study was approved under Caltech IRB #20-1026. All adult participants provided written informed consent; all minor participants provided verbal assent accompanied by written permission from a legal guardian. Children ages 8-17 years old additionally provided written assent.

##### Additional Details about RT-qPCR Testing

Briefly, each day, participants completed an online symptom survey, then self-collected saliva, then anteriornares swab, then posterior oropharyngeal (throat) swab specimens for RT-qPCR testing. Extraction and RT-qPCR was performed at Pangea Laboratories using the FDA-authorized Quick SARS-CoV-2 RT-qPCR Kit.^30^ This assay has a reported LOD of 250 copies/mL of sample, which we also verified prior to study initiation.^25^ Details of the quantification of viral load were described previously.^25^

Viral sequencing and variant determination were also performed at Pangea; full methods previously described.^25^ Extraction, RT-qPCR, and sequencing operators and supervisors at Pangea Laboratory were blinded to which participant a sample originated from, as well as the infection status and Ag-RDT results of all participants.

### Supplementary Analyses

#### Discordance in Participant Interpretation of Antigen Test Results

Participants interpreted and reported their own antigen test results (positive, negative, or invalid), and photographed their test strips immediately. In the event of an invalid result, study coordinators contacted participants to request they immediately take an additional test; invalid results were replaced with subsequent valid results, when applicable. Participants recorded their test results and uploaded photos of the test strips to a secure REDCap server immediately after testing. All photographs were inspected by at least two study coordinators blinded to RT-qPCR results. Results as reported by the participants were analyzed and reported here. In 2.5% of antigen tests (56 of 2,153 tests), a pink (positive) test line was visible to two study coordinators in photographs uploaded, but the result was reported as negative by the participant. In most cases the pink lines were faint and may have been overlooked by the participants. It is also possible that in some cases the test was photographed late; per the manufacturer’s guidance, the test result is only valid at the 10-min mark. One participant with a dark pink line was queried and reported poor close-range vision; this participant had a housemate help with all further interpretations. In one case from one participant, an invalid result was reported, but a blue control line was visible to two study coordinators. In this manuscript, we used the participants’ interpretations in all analyses. Although 2.5% of all rapid antigen test results had discordant interpretations, 14% (33 of 228) participants had a discordant interpretation; this discordance underlines that user error can affect sensitivity of these at-home tests in real-world settings.

#### Faulty Antigen Test Lot

In mid-January 2022 we observed that two asymptomatic participants had consecutive positive antigen test results, but negative results by RT-qPCR in all three specimen types tested. Further investigation revealed that the most recently taken false positives from these two participants were from the same antigen strip lot (Quidel QuickVue At-Home OTC COVID-19 Test #152000). A third participant (**Fig 4D**) also had a single false-positive test from this lot the same week. This lot was immediately pulled from circulation in the study, and reported to the manufacturer and to the FDA (via a MedWatch Voluntary Report). Following an IRB amendment, participants began photographing the antigen test strip lot number visible when they reported their Ag-RDT results. Known test results from this faulty lot were marked as invalid and excluded from analysis (**Fig 2**). In one of the 17 participants enrolled during the early period of infection (**Fig 4D**), the antigen test result from this lot is noted with a “?” on his plot, and the datapoint was excluded from subsequent analyses. An investigation of the high rate of false positives was investigated further in a laboratory study using antigen test buffer and commercial nasal fluid from healthy human donors. Full details of that investigation have been reported separately.^1^

In the participant in **Fig 4D**, we continued to observe consistent false-positive Ag-RDTs; with a variety of antigen lots. The participant also tested positive by the Quidel Ag-RDT while testing negative on an iHealth rapid antigen test taken outside of the study on the final day of sampling. This participant tested positive by the Quidel Ag-RDT even >30 days after his first detectable viral load, and when viral load was undetectable by RT-qPCR in all three specimen types. These antigen test strips were not from the lot that yielded consistently false-positive results. The reason for this participant’s string of false positives remains unknown.

We observed a negative percent agreement (NPA) of 97% (1,343) antigen negative results of 1,385 ANS RT-qPCR negative results. This is slightly lower than the NPA of 99.2% (95% CI 97.2-99.8%) observed by the Ag-RDT manufacturer.^32^ This decrease may be due to the inclusion of additional results from this faulty lot for which we were not able to collect test strip lot information.

**Table S1.** Demographic and Medical Information for the Participants Shown in Fig 4. SARS-CoV-2 variant was determined by ANS swab in all cases except individual (B) who had low ANS viral loads so viral load was sequenced from a throat swab. The variant for participant (I) is inferred from the household index case. Summary information can be found in Table S2.

| Fig 3 panel | Status on enrollment |  |  |  | Months since vaccine |  |  | Active Medications | Medical conditions | Gender | Age range (in years) | Race | Ethnicity | SARS-CoV-2 Variant |
| --- | --- | --- | --- | --- | --- | --- | --- | --- | --- | --- | --- | --- | --- | --- |
|  | Saliva PCR | Throat PCR | Nasal PCR | Nasal antigen | 1st dose | 2nd dose | 3rd dose |  |  |  |  |  |  |  |
| (A) | neg | neg | neg | neg | 9 [M] | 8 [M] | <2 [M] | n/a | n/a | male | 40-49 | White | not Hispanic | Omicron BA.1.1 |
| (B) | neg | neg | neg | neg | 11 [JJ] | 3 [P] | none | PPI, vitamin/ supplement | obesity, GI condition, anxiety or depression | female | 30-39 | White | not Hispanic | Omicron BA.1.1 |
| (C) | inc | neg | neg | neg | <1 [P] | none | none | acetaminophen | n/a | male | 6-11 | Multiple Races | not Hispanic | Omicron BA.1.1 |
| (D) | neg | neg | neg | neg | 10 [M] | 9 [M] | 2 [M] | none | obesity | male | 30-39 | Asian or Pacific Islander | not Hispanic | Omicron BA.1.1 |
| (E) | neg | neg | neg | neg | >11 [P] | <10 [P] | <3 [P] | allergy medication; acetaminophen, antihistamine, dextromethorphan, phenylephrine HCl, doxylamine | obesity | female | 30-39 | White | Hispanic | Omicron BA.1 |
| (F) | neg | neg | neg | neg | 10 [P] | 9 [P] | none | vitamin/ supplement | n/a | female | 18-29 | White | not Hispanic | Omicron BA.1.1 |
| (G) | neg | neg | neg | neg | <2 [P] | <1 [P] | none | vitamin/ supplement | n/a | male | 6-11 | White | not Hispanic | Omicron BA.1.1 |
| (H) | neg | neg | neg | neg | 10 [M] | 9 [M] | 2 [M] | vitamin/ supplement | n/a | female | 40-49 | White | not Hispanic | Omicron BA.1.1 |
| (I) | neg | neg | neg | neg | 10 [P] | 9 [P] | none | antibiotic, vitamin/ supplement | obesity | male | 18-29 | White | Hispanic | Omicron BA.1.1 (index case) |
| (J) | pos | pos | inc | neg | 9 [M] | 8 [M] | <2 [M] | vitamin/ supplement | anxiety or depression | female | 40-49 | White | not Hispanic | Omicron BA.1.1 |
| (K) | pos | pos | inc | neg | 9.5 [M] | 8.5 [M] | 0.5 [P] | NSAID | n/a | male | 40-49 | White | not Hispanic | Omicron BA.1.1 |
| (L) | pos | pos | pos | neg | 11 [P] | 10 [P] | 2 [P] | allergy medication, diabetes medication, cholesterol medication | diabetes, high blood pressure, obesity, asthma, sleep apnea, GI condition | female | 50-59 | Multiple Races | not Hispanic | Omicron BA.1.1 |
| (M) | pos | pos | neg | neg | 10 [M] | 9 [M] | 2 [M] | SSRI | oveweight, anxiety or depression | male | 50-59 | White | not Hispanic | Omicron BA.1.1 |
| (N) | pos | neg | pos | neg | 5 [P] | 4[P] | none | none | n/a | female | 12-17 | White | not Hispanic | Omicron BA.1.1 |
| (O) | pos | pos | pos | neg | 10 [P] | 9 [P] | 1 [P] | vitamin/ supplement | anxiety or depression | female | 40-49 | White | not Hispanic | Omicron BA.1.1 |
| (P) | pos | pos | pos | neg | 13 [P] | 12 [P] | 3.5 [P] | none | n/a | male | 18-29 | Asian | not Hispanic | Omicron BA.1.1 |
| (Q) | pos | pos | pos | neg | 9[P] | 8[P] | <0.5 [P] | acetaminophen, antihistamine, dextromethorphan, phenylephrine HCl, doxylamine | obesity | female | 12-17 | White | Hispanic | Omicron BA.1 |

\* Months from vaccine date are given relative to enrollment date

**Table S2.** Demographics of the 17-participant cohort shown in Fig 4. Additional detailed information on each participant can be found in Table S1.

| **<u>Sex\*</u>** |  |  |
| --- | --- | --- |
| Male | 8 | 47.1% |
| Female | 9 | 52.9% |
| **<u>Age</u>** |  |  |
| 6-11 | 2 | 11.8% |
| 12-17 | 2 | 11.8% |
| 18-29 | 3 | 17.6% |
| 30-39 | 3 | 17.6% |
| 40-49 | 5 | 29.4% |
| 50-59 | 2 | 11.8% |
| **<u>Race</u>** |  |  |
| White | 13 | 76.5% |
| Asian or Pacific Islander | 2 | 11.8% |
| Multiple Races | 2 | 11.8% |
| **<u>Ethnicity</u>** |  |  |
| Hispanic | 3 | 17.6% |
| Non-Hispanic | 14 | 82.4% |
| **<u>Tobacco Smoker or Vape User History</u>** |  |  |
| Current | 0 | 0.0% |
| Former | 2 | 11.8% |
| Never | 15 | 88.2% |
| **<u>Active Medications and Supplements</u>** |  |  |
| Vitamins/Supplements | 7 | 41.2% |
| Acetaminophen/NSAIDs | 4 | 23.5% |
| Allergy medications/Antihistamines | 3 | 17.6% |
| Antibiotics/Antivirals | 1 | 5.9% |
| **<u>Medical Comorbidities</u>** |  |  |
| Asthma | 1 | 5.9% |
| Anxiety or Depression | 3 | 17.6% |
| Diabetes | 1 | 5.9% |
| Overweight/Obesity | 7 | 41.2% |
| GI condition | 2 | 11.8% |
| **<u>SARS-CoV-2 Vaccination Status</u>** |  |  |
| Partially Vaccinated | 1 | 5.9% |
| Completed Vaccination | 5 | 29.4% |
| Fully vaccinated and boosted | 11 | 64.7% |
| No SARS-CoV-2 vaccines reported | 0 | 0.0% |

\*Participants were asked to report both sex at birth and current gender identity; all participants in this cohort responded cis-gender identities to sex at birth

**Figure S1.**
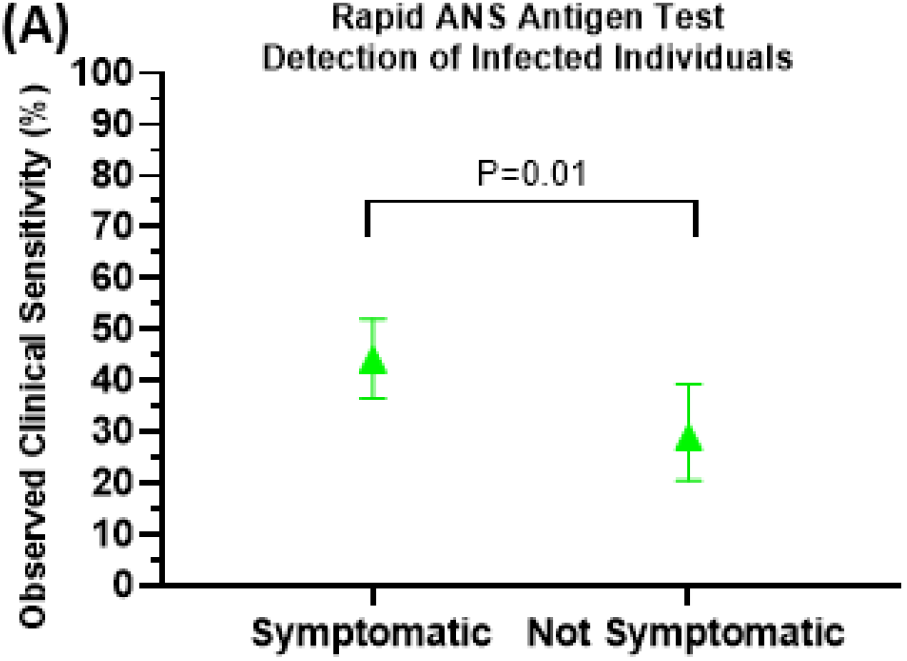
Relationship Between Symptoms and Viral Load. The observed clinical sensitivity of the rapid antigen test to detect infection is plotted for timepoints when the cohort of 17 participants enrolled early in the course of the infection either reported at least one symptom (Symptomatic) or did not report any symptoms (Not Symptomatic). An upper-tailed Fished exact test was performed to determine whether Ag-RDT performance at symptomatic timepoints was significantly higher than timepoints when participants experienced no symptoms.

**Figure S2.**
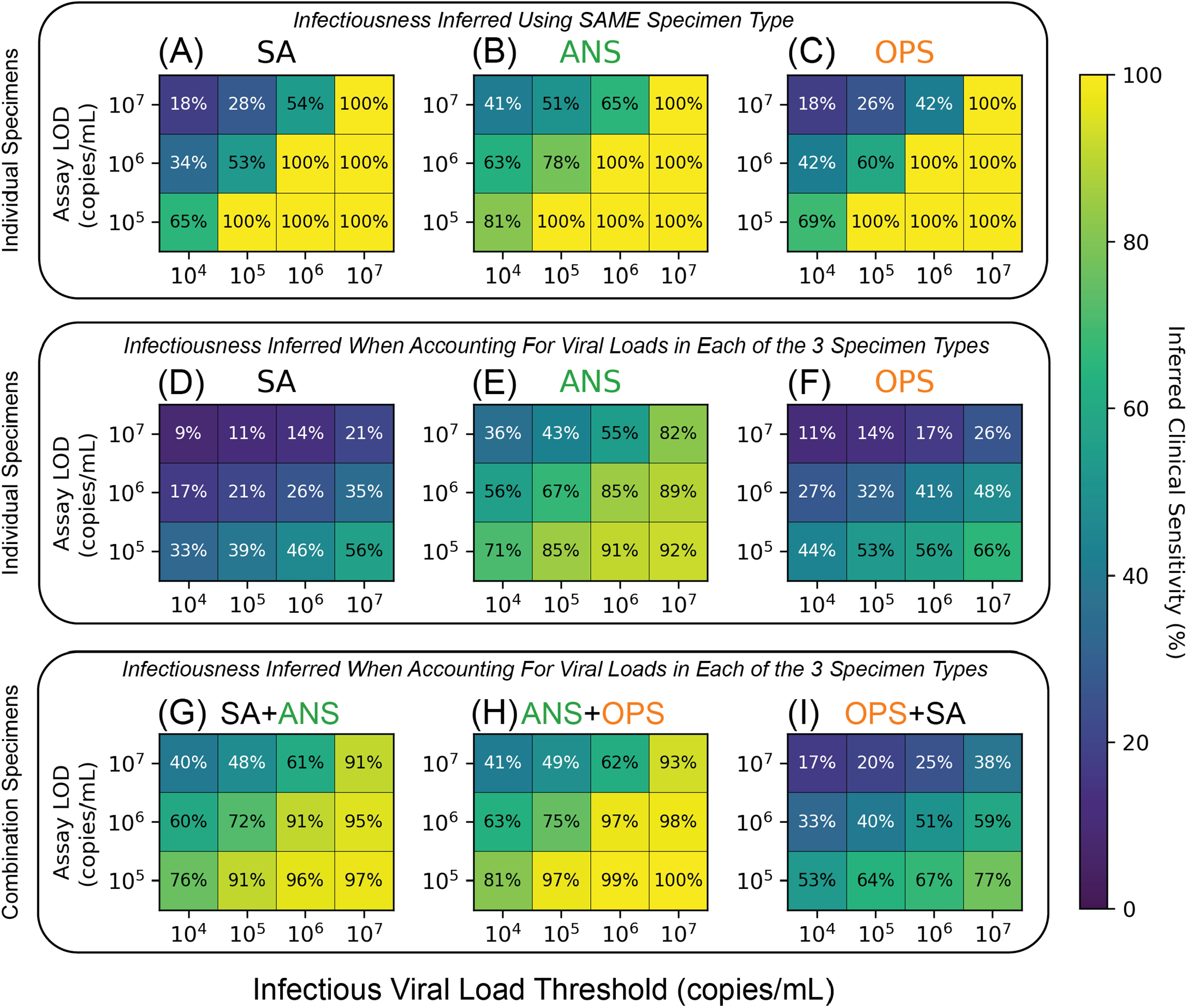
Effect of Test LOD and Infectious Viral-Load Threshold (IVLT) on Inferred Clinical Sensitivity of Contrived Specimen Combinations. Clinical sensitivities of assays with varying LOD and IVLT for single specimen types (A-F) and contrived combination specimen types (G-I). Samples were deemed infectious if its own viral load surpassed the IVLT (A-C), or if the viral load any sample collected from the same individual at the same timepoint surpassed the IVLT (D-I). Contrived combination specimens (G-I) were calculated by taking the max viral load over the two specified specimen types.

## AUTHOR CONTRIBUTIONS (listed alphabetically by last name)

Reid Akana (RA): Collaborated with AVW in creating digital participant symptom surveys; assisted with data quality control/curation with NS, HD, SC; created current laboratory information management system (LIMS) for specimen logging and tracking. Creation of iOS application for sample logging/tracking. Configured an SQL database for data storage. Created an Apache server and websites to view study data. Configured FTPS server to catalog PCR data. Wrote a Python package to access study data. Trained study coordinators on SQL. Troubleshooting and QC of LIMS. Made Figures 3, 5, S2, S3, S4, S5. Wrote and edited the manuscript with AVW and NS.

Alyssa M. Carter (AMC): Assisted with the inventory and archiving of >6,000 samples at Caltech; coordinated shipment of samples to Caltech with AER and JRBR; assisted with procurement of antigen tests; assisted with organizing volunteers and making participant kits; assisted AER in developing and implementing QC for participant kits. Led the in-lab investigation of antigen false-positive results; designed and performed experiments for lot analysis of the Quidel QuickVue At-Home Covid-19 tests. Provided feedback and edited the manuscript.

Yap Ching Chew (YCC): Primary liaison with Caltech team. Prepared and provided Zymo SafeCollect kits and related materials to Caltech team. Supervised the extraction, PCR, and QC teams at Pangea Laboratory. Sent PCR results daily to Caltech team. Arranged for Pangea team to perform viral-variant sequencing on selected samples; reported results and provided sequencing files.

Saharai Caldera (SC): Study coordinator; recruited, enrolled and maintained study participants with NS and HD; study-data quality control, curation and archiving with RA, NS, HD and MKK; supplies acquisition with AER, NS, HD and MKK.

Hannah Davich (HD): Lead study coordinator; co-wrote participant informational sheets with NS; developed recruitment strategies and did outreach with NS; participant kit creation and co-coordinated kit-making by volunteers with AER; recruited, enrolled and maintained study participants with NS and SC; managed the study-coordinator inventory; study-data quality control, curation and archiving with RA, NS, SC and MKK; supplies acquisition with AER, NS, SC and MKK.

Matthew Feaster (MF): Co-investigator; collaborated with AVW, MMC, NS, YG, RFI on study design and recruitment strategies; provided guidance and expertise on SARS-CoV-2 epidemiology and local trends.

Ying-Ying Goh (Y-YG): Co-investigator; collaborated with AVW, MMC, NS, MF, RFI on study design and recruitment strategies; provided guidance and expertise on SARS-CoV-2 epidemiology and local trends.

Rustem F. Ismagilov (RFI): Principal investigator; collaborated with AVW, MMC, NS, MF, YYG on study design and recruitment strategies; provided leadership, technical guidance, and oversight of all analyses; was responsible for obtaining the primary funding for the study.

Mi Kyung Kim (MKK): Study coordinator (part-time); maintaining participants with NS, HD, and SC; study-data quality control, curation and archiving with RA, NS, SC and HD; supplies acquisition with AER, NS, SC and HD; collected contact info for local health centers for recruitment outreach; assembled Table S1 with NS.

John Raymond B. Reyna (JRBR): Organized sample labeling and short-term storage of all samples at Pangea Laboratories. Arranged shipment of all samples to Caltech team. Assisted with processing of the specimens.

Anna E. Romano (AER): Co-coordinated kit-making by volunteers with HD; implemented QC process for kit-making; participated in kit making; managed logistics for the inventory and archiving of >6,000 samples at Caltech; supplies acquisition with HD, NS, SC and MKK; assisted with securing funding; compiled antigen lot data to assist false-positive antigen test investigation; organized and performed QC on sequencing data. Provided feedback and edited the manuscript.

Natasha Shelby (NS): Study administrator; collaborated with AVW, RFI, YG, MF on initial study design and recruitment strategies; co-wrote IRB protocol and informed consent with AVW; co-wrote enrollment questionnaire and post-study questionnaire with AVW; initiated the collaboration with Zymo and served as primary liaison throughout study; reviewed pilot sampling data and amended instructional sheets/graphics for specimen collections in collaboration with Zymo; co-wrote participant informational sheets with HD; hired, trained, and supervised the study-coordinator team; developed recruitment strategies and did outreach with HD; recruited, enrolled and maintained study participants with HD and SC; co-developed participant keep/drop criteria with AVW; performed the daily upload, review, and QC of PCR data received from Zymo; made the daily keep/drop decisions based on viral-load trajectories in each household; made all phone calls to alert presumptive positives of their status and provide resources; study-data quality control, curation and archiving with RA, HD, SC and MKK; organized archiving of all participant data and antigen-test photographs; supplies acquisition with AER, HD, SC and MKK; assisted with securing funding; managed the overall study budget; assembled Fig 1 with AVW; assembled Table S1 with MKK; made Fig 4 with AVW; managed citations and reference library; verified the underlying data with AVW and RA; co-wrote and edited the manuscript with AVW and RA.

Matt Thomson (MT): Assisted with statistical approach and analyses.

Colten Tognazzini (CT): Coordinated the recruitment efforts at PPHD with case investigators and contact tracers; provided guidance and expertise on SARS-CoV-2 epidemiology and local trends.

Alexander Viloria Winnett (AVW): Collaborated with NS, RFI, YG, MF on initial study design and recruitment strategies; co-wrote IRB protocol and informed consent with NS; co-wrote enrollment questionnaire and post-study questionnaire with NS; co-developed participant keep/drop criteria with NS; funding acquisition; designed and coordinated LOD validation experiments; selected and prepared specimen for viral-variant sequencing with NS, YC, and AER; assisted with the inventory and archiving of >6,000 specimen at Caltech with AER and AMC; minor role supporting outreach by HD and NS; minor role supporting kit-making by AER, HD and AMC; verified the underlying data with NS and RA; major contributor to reference organization and selection; assembled Fig 1 with NS; made Fig 4 with NS; performed analysis and prepared Fig 2, Fig 6, Fig 7, Fig S1, and Table S2. Co-wrote and edited the manuscript with NS and RA.

Taikun Yamada (TY): Performed the RT-qPCR COVID-19 testing at Pangea Laboratory.

## References

1. WHO. Antigen-detection in the diagnosis of SARS-CoV-2 infection [interim guidance]. 2021. https://www.who.int/publications/i/item/antigen-detection-in-the-diagnosis-of-sars-cov-2infection-using-rapid-immunoassays.

2. PATH. Global Availability of COVID-19 Diagnostic Tests. 2022. https://www.path.org/programs/diagnostics/covid-dashboard-global-availability-covid-19-diagnostic-tests/.

3. European Centre for Disease Prevention and Control. Diagnostic Testing and Screening for SARS-CoV-2. 2022. https://www.ecdc.europa.eu/en/covid-19/latest-evidence/diagnostic-testing.

4. National Health Service. How to use an NHS rapid lateral flow test for coronavirus (COVID-19). 2021. https://www.nhs.uk/conditions/coronavirus-covid-19/testing/how-to-do-a-test-at-home-or-at-a-test-site/how-to-do-a-rapid-lateral-flow-test/.

5. Drain PK. Rapid Diagnostic Testing for SARS-CoV-2. N Engl J Med 2022; 386(3): 264–72.

6. Rader B, Gertz A, Iuliano AD, et al. Use of At-Home COVID-19 Tests - United States, August 23, 2021-March 12, 2022. MMWR Morb Mortal Wkly Rep 2022; 71(13): 489–94.

7. Hayden MK, Hanson KE, Englund JA, et al. The Infectious Diseases Society of America Guidelines on the Diagnosis of COVID-19: Antigen Testing. IDSA 2022; v.2 updated 12/20/2022.

8. Procop GW, Kadkhoda K, Rhoads DD, Gordon SG, Reddy AJ. Home testing for COVID-19: Benefits and limitations. Cleve Clin J Med 2021.

9. Han AX, Girdwood SJ, Khan S, et al. Strategies for Using Antigen Rapid Diagnostic Tests to Reduce Transmission of Severe Acute Respiratory Syndrome Coronavirus 2 (SARS-CoV-2) in Low- and Middle-Income Countries: A Mathematical Modelling Study Applied to Zambia. Clin Infect Dis 2022: ciac814.

10. Hardick J, Gallagher N, Sachithanandham J, et al. Evaluation of Four Point of Care (POC) Antigen Assays for the Detection of the SARS-CoV-2 Variant Omicron. Microbiol Spectr 2022: e0102522.

11. FDA. In Vitro Diagnostic EUAs - Other Tests for SARS-CoV-2. 2022. https://www.fda.gov/medical-devices/coronavirus-disease-2019-covid-19-emergency-use-authorizations-medical-devices/in-vitro-diagnostics-euas-other-tests-sars-cov-2.

12. Cubas-Atienzar AI, Kontogianni K, Edwards T, et al. Limit of detection in different matrices of 19 commercially available rapid antigen tests for the detection of SARS-CoV-2. Sci Rep 2021; 11(1): 18313.

13. FDA. Omicron Variant: Impact on Antigen Diagnostic Tests (As of 12/28/2021). 2021. https://www.fda.gov/medical-devices/coronavirus-covid-19-and-medical-devices/sars-cov-2-viral-mutations-impact-covid-19-tests#omicronvariantimpact.

14. Guglielmi G. Nature News Feature: Rapid coronavirus tests: a guide for the perplexed. 2021. https://www.nature.com/articles/d41586-021-00332-4.

15. Mercer T, Almond N, Crone MA, et al. The Coronavirus Standards Working Group’s roadmap for improved population testing. Nat Biotechnol 2022; 40(11): 1563–8.

16. Stanley S, Hamel Donald J, Wolf Ian D, et al. Limit of Detection for Rapid Antigen Testing of the SARS-CoV-2 Omicron and Delta Variants of Concern Using Live-Virus Culture. J Clin Microbiol 2022; 60(5): e00140–22.

17. Deerain J, Druce J, Tran T, et al. Assessment of the Analytical Sensitivity of 10 Lateral Flow Devices against the SARS-CoV-2 Omicron Variant. J Clin Microbiol 2022; 60(2): e0247921–e.

18. Lee Rose A, Herigon Joshua C, Benedetti A, Pollock Nira R, Denkinger Claudia M, Humphries Romney M. Performance of Saliva, Oropharyngeal Swabs, and Nasal Swabs for SARS-CoV-2 Molecular Detection: a Systematic Review and Meta-analysis. J Clin Microbiol; 59(5): e02881–20.

19. Patriquin G, Davidson RJ, Hatchette TF, et al. Generation of False-Positive SARS-CoV-2 Antigen Results with Testing Conditions outside Manufacturer Recommendations: A Scientific Approach to Pandemic Misinformation. Microbiol Spectr 2021; 9(2): e0068321.

20. Torres I, Poujois S, Albert E, Colomina J, Navarro D. Real-life evaluation of a rapid antigen test (Panbio COVID-19 Ag Rapid Test Device) for SARS-CoV-2 detection in asymptomatic close contacts of COVID-19 patients. medRxiv 2020: 2020.12.01.20241562.

21. Pray IW, Ford L, Cole D, et al. Performance of an Antigen-Based Test for Asymptomatic and Symptomatic SARS-CoV-2 Testing at Two University Campuses - Wisconsin, September-October 2020. MMWR Morb Mortal Wkly Rep 2021; 69(5152): 1642–7.

22. Soni A, Herbert C, Lin H, et al. Performance of Screening for SARS-CoV-2 using Rapid Antigen Tests to Detect Incidence of Symptomatic and Asymptomatic SARS-CoV-2 Infection: findings from the Test Us at Home prospective cohort study. medRxiv 2022: 2022.08.05.22278466.

23. Tinker SC, Prince-Guerra JL, Vermandere K, et al. Evaluation of self-administered antigen testing in a college setting. Virol J 2022; 19(1): 202.

24. California Department of Public Health. Testing Framework for K–12 Schools for the 2022–2023 School Year. 2022. https://www.cdph.ca.gov/Programs/CID/DCDC/Pages/COVID-19/Testing-Framework-for-K-12-Schools-for-the-2022-2023-School-Year.aspx.

25. Holmdahl I, Kahn R, Hay J, Buckee CO, Mina M. Estimation of Transmission of COVID-19 in Simulated Nursing Homes With Frequent Testing and Immunity-Based Staffing. JAMA Network Open 2021; 4(5): e2110071.

26. Larremore DB, Wilder B, Lester E, et al. Test sensitivity is secondary to frequency and turnaround time for COVID-19 screening. Science Advances 2021; 7(1): eabd5393.

27. Mina MJ, Parker R, Larremore DB. Rethinking Covid-19 Test Sensitivity — A Strategy for Containment. N Engl J Med 2020.

28. Tom MR, Mina MJ. To Interpret the SARS-CoV-2 Test, Consider the Cycle Threshold Value. Clin Infect Dis 2020; 71(16): 2252–4.

29. Bullard J, Dust K, Funk D, et al. Predicting Infectious Severe Acute Respiratory Syndrome Coronavirus 2 From Diagnostic Samples. Clin Infect Dis 2020; 71(10): 2663–6.

30. Marc A, Kerioui M, Blanquart F, et al. Quantifying the relationship between SARS-CoV-2 viral load and infectiousness. eLife 2021; 10: e69302.

31. Walsh KA, Jordan K, Clyne B, et al. SARS-CoV-2 detection, viral load and infectivity over the course of an infection. The Journal of infection 2020; 81(3): 357–71.

32. van Kampen JJA, van de Vijver DAMC, Fraaij PLA, et al. Duration and key determinants of infectious virus shedding in hospitalized patients with coronavirus disease-2019 (COVID-19). Nature communications 2021; 12(1): 267.

33. Perera R, Tso E, Tsang OTY, et al. SARS-CoV-2 Virus Culture and Subgenomic RNA for Respiratory Specimens from Patients with Mild Coronavirus Disease. Emerg Infect Dis 2020; 26(11): 2701–4.

34. Pickering S, Batra R, Merrick B, et al. Comparative performance of SARS-CoV-2 lateral flow antigen tests and association with detection of infectious virus in clinical specimens: a single-centre laboratory evaluation study. Lancet Microbe 2021; 2(9): e461–e71.

35. Perera RAPM, Tso E, Tsang OTY, et al. SARS-CoV-2 virus culture from the upper respiratory tract: Correlation with viral load, subgenomic viral RNA and duration of illness. medRxiv 2020: 2020.07.08.20148783.

36. L’Huillier AG, Torriani G, Pigny F, Kaiser L, Eckerle I. Shedding of infectious SARS-CoV-2 in symptomatic neonates, children and adolescents. medRxiv 2020: 2020.04.27.20076778.

37. Jones Terry C, Biele G, Mühlemann B, et al. Estimating infectiousness throughout SARS-CoV-2 infection course. Science 2021; 373(6551): eabi5273.

38. Quicke K, Gallichote E, Sexton N, et al. Longitudinal Surveillance for SARS-CoV-2 RNA Among Asymptomatic Staff in Five Colorado Skilled Nursing Facilities: Epidemiologic, Virologic and Sequence Analysis. medRxiv 2020: 2020.06.08.20125989.

39. Puhach O, Adea K, Hulo N, et al. Infectious viral load in unvaccinated and vaccinated patients infected with SARS-CoV-2 WT, Delta and Omicron. medRxiv 2022: 2022.01.10.22269010.

40. Bal A, Brengel-Pesce K, Gaymard A, et al. Clinical and microbiological assessments of COVID-19 in healthcare workers: a prospective longitudinal study. medRxiv 2020: 2020.11.04.20225862.

41. Ke R, Martinez PP, Smith RL, et al. Daily longitudinal sampling of SARS-CoV-2 infection reveals substantial heterogeneity in infectiousness. Nature Microbiology 2022; 7(5): 640–52.

42. Boucau J, Marino C, Regan J, et al. Duration of Shedding of Culturable Virus in SARS-CoV-2 Omicron (BA.1) Infection. N Engl J Med 2022.

43. Killingley B, Mann AJ, Kalinova M, et al. Safety, tolerability and viral kinetics during SARS-CoV-2 human challenge in young adults. Nat Med 2022; 28(5): 1031–41.

44. Wölfel R, Corman VM, Guggemos W, et al. Virological assessment of hospitalized patients with COVID-2019. Nature 2020; 581(7809): 465–9.

45. Rhee C, Kanjilal S, Baker M, Klompas M. Duration of Severe Acute Respiratory Syndrome Coronavirus 2 (SARS-CoV-2) Infectivity: When Is It Safe to Discontinue Isolation? Clin Infect Dis 2021; 72(8): 1467–74.

46. Pekosz A, Parvu V, Li M, et al. Antigen-Based Testing but Not Real-Time Polymerase Chain Reaction Correlates With Severe Acute Respiratory Syndrome Coronavirus 2 Viral Culture. Clin Infect Dis 2021; 73(9): e2861–e6.

47. Cevik M, Tate M, Lloyd O, Maraolo AE, Schafers J, Ho A. SARS-CoV-2, SARS-CoV, and MERS-CoV viral load dynamics, duration of viral shedding, and infectiousness: a systematic review and meta-analysis. Lancet Microbe 2021; 2(1): e13–e22.

48. Kirby JE, Riedel S, Dutta S, et al. SARS-CoV-2 antigen tests predict infectivity based on viral culture: comparison of antigen, PCR viral load, and viral culture testing on a large sample cohort. Clin Microbiol Infect 2023; 29(1): 94–100.

49. Ke R, Martinez PP, Smith RL, et al. Longitudinal analysis of SARS-CoV-2 vaccine breakthrough infections reveal limited infectious virus shedding and restricted tissue distribution. Open Forum Infectious Diseases 2022: ofac192.

50. Bonenfant G, Deyoe JE, Wong T, et al. Surveillance and Correlation of Severe Acute Respiratory Syndrome Coronavirus 2 Viral RNA, Antigen, Virus Isolation, and Self-Reported Symptoms in a Longitudinal Study With Daily Sampling. Clin Infect Dis 2022; 75(10): 1698–705.

51. Adamson B, Sikka R, Wyllie AL, Premsrirut P. Discordant SARS-CoV-2 PCR and Rapid Antigen Test Results When Infectious: A December 2021 Occupational Case Series. medRxiv 2022: 2022.01.04.22268770.

52. Savela ES, Viloria Winnett A, Romano Anna E, et al. Quantitative SARS-CoV-2 viral-load curves in paired saliva and nasal swabs inform appropriate respiratory sampling site and analytical test sensitivity required for earliest viral detection. J Clin Microbiol 2021; 0(ja): JCM.01785-21.

53. Winnett A, Cooper MM, Shelby N, et al. SARS-CoV-2 Viral Load in Saliva Rises Gradually and to Moderate Levels in Some Humans. medRxiv 2020: 2020.12.09.20239467.

54. Lai J, German J, Hong F, et al. Comparison of Saliva and Mid-Turbinate Swabs for Detection of COVID-19. medRxiv 2022: 2021.12.01.21267147.

55. Viloria Winnett A, Akana R, Shelby N, et al. SARS-CoV-2 exhibits extreme differences in early viral loads among specimen types suggesting improved detection of pre-infectious and infectious individuals using combination specimen types. medRxiv 2022: MEDRXIV/2022/277113.

56. Stankiewicz Karita HC, Dong TQ, Johnston C, et al. Trajectory of Viral RNA Load Among Persons With Incident SARS-CoV-2 G614 Infection (Wuhan Strain) in Association With COVID-19 Symptom Onset and Severity. JAMA Network Open 2022; 5(1): e2142796–e.

57. Kissler SM, Fauver JR, Mack C, et al. Viral dynamics of acute SARS-CoV-2 infection and applications to diagnostic and public health strategies. PLoS Biol 2021; 19(7): e3001333–e.

58. Kissler SM, Fauver JR, Mack C, et al. Viral Dynamics of SARS-CoV-2 Variants in Vaccinated and Unvaccinated Persons. N Engl J Med 2021; 385(26): 2489–91.

59. Moreno GK, Braun KM, Pray IW, et al. Severe Acute Respiratory Syndrome Coronavirus 2 Transmission in Intercollegiate Athletics Not Fully Mitigated With Daily Antigen Testing. Clin Infect Dis 2021; 73(Supplement_1): S45–S53.

60. Li R, Pei S, Chen B, et al. Substantial undocumented infection facilitates the rapid dissemination of novel coronavirus (SARS-CoV-2). Science 2020; 368(6490): 489–93.

61. Robertson LS. Did people’s behavior after receiving negative COVID-19 tests contribute to the spread? Journal of public health (Oxford, England) 2021; 43(2): 270–3.

62. FDA. Emergency Use Authorization (EUA) information, and list of all current EUAs. 2020. https://www.fda.gov/emergency-preparedness-and-response/mcm-legal-regulatory-and-policy-framework/emergency-use-authorization#2019-ncov.

63. Chu VT, Schwartz NG, Donnelly MAP, et al. Comparison of Home Antigen Testing With RT-PCR and Viral Culture During the Course of SARS-CoV-2 Infection. JAMA Internal Medicine 2022; 182(7): 701–9.

64. Korenkov M, Poopalasingam N, Madler M, et al. Evaluation of a Rapid Antigen Test To Detect SARS-CoV-2 Infection and Identify Potentially Infectious Individuals. J Clin Microbiol 2021; 59(9): e00896–21.

65. Smith RL, Gibson LL, Martinez PP, et al. Longitudinal Assessment of Diagnostic Test Performance Over the Course of Acute SARS-CoV-2 Infection. The Journal of Infectious Diseases 2021; 224(6): 976–82.

66. Bouton TC, Atarere J, Turcinovic J, et al. Viral Dynamics of Omicron and Delta Severe Acute Respiratory Syndrome Coronavirus 2 (SARS-CoV-2) Variants With Implications for Timing of Release from Isolation: A Longitudinal Cohort Study. Clin Infect Dis 2022: ciac510.

67. Kohmer N, Toptan T, Pallas C, et al. The Comparative Clinical Performance of Four SARS-CoV-2 Rapid Antigen Tests and Their Correlation to Infectivity In Vitro. Journal of clinical medicine 2021; 10(2).

68. Steinlin-Schopfer J, Barbani MT, Kamgang R, Zwahlen M, Suter-Riniker F, Dijkman R. Evaluation of the Roche antigen rapid test and a cell culture-based assay compared to rRT-PCR for the detection of SARS-CoV-2: A contribution to the discussion about SARS-CoV-2 diagnostic tests and contagiousness. Journal of clinical virology plus 2021; 1(1): 100020.

69. Ford L, Lee C, Pray IW, et al. Epidemiologic Characteristics Associated With Severe Acute Respiratory Syndrome Coronavirus 2 (SARS-CoV-2) Antigen-Based Test Results, Real-Time Reverse Transcription Polymerase Chain Reaction (rRT-PCR) Cycle Threshold Values, Subgenomic RNA, and Viral Culture Results From University Testing. Clin Infect Dis 2021; 73(6): e1348–e55.

70. Shah MM, Salvatore PP, Ford L, et al. Performance of Repeat BinaxNOW Severe Acute Respiratory Syndrome Coronavirus 2 Antigen Testing in a Community Setting, Wisconsin, November 2020–December 2020. Clin Infect Dis 2021; 73(Supplement_1): S54–S7.

71. Prince-Guerra JL, Almendares O, Nolen LD, et al. Evaluation of Abbott BinaxNOW Rapid Antigen Test for SARS-CoV-2 Infection at Two Community-Based Testing Sites - Pima County, Arizona, November 3-17, 2020. MMWR Morb Mortal Wkly Rep 2021; 70(3): 100–5.

72. Pilarowski G, Marquez C, Rubio L, et al. Field Performance and Public Health Response Using the BinaxNOWTM Rapid Severe Acute Respiratory Syndrome Coronavirus 2 (SARS-CoV-2) Antigen Detection Assay During Community-Based Testing. Clin Infect Dis 2021; 73(9): e3098–e101.

73. Pollock NR, Jacobs JR, Tran K, et al. Performance and Implementation Evaluation of the Abbott BinaxNOW Rapid Antigen Test in a High-Throughput Drive-Through Community Testing Site in Massachusetts. J Clin Microbiol 2021; 59(5).

74. FDA. Quidel QuickVue At-Home OTC COVID-19 Test - Instructions for Use (IFU). 2021. https://www.fda.gov/media/147265/download

75. Quidel. Quidel QuickVue At-Home OTC COVID-19 Test - User Instructions. 2021. https://quickvueathome.com/wp-content/uploads/2021/05/EF1479701EN00_QV_At-Home_Web_Instructions_English_052421.pdf.

76. Kepczynski CM, Genigeski JA, Koski RR, Bernknopf AC, Konieczny AM, Klepser ME. A systematic review comparing at-home diagnostic tests for SARS-CoV-2: Key points for pharmacy practice, including regulatory information. J Am Pharm Assoc (2003) 2021; 61(6): 666–77.e2.

77. Bourassa L, Perchetti GA, Phung Q, et al. A SARS-CoV-2 Nucleocapsid Variant that Affects Antigen Test Performance. J Clin Virol 2021; 141: 104900.

78. CLSI. EP12-A2 User Protocol for Evaluation of Qualitative Test Performance, 2nd edition; 2008.

79. FDA. At-Home COVID-19 Antigen Tests-Take Steps to Reduce Your Risk of False Negative: FDA Safety Communication. 2022. https://www.fda.gov/medical-devices/safety-communications/home-covid-19-antigen-tests-take-steps-reduce-your-risk-false-negative-fda-safety-communication?utm_medium=email&utm_source=govdelivery.

80. CDC. Guidance for Antigen Testing for SARS-CoV-2 for Healthcare Providers Testing Individuals in the Community. 2022. https://www.cdc.gov/coronavirus/2019-ncov/lab/resources/antigen-tests-guidelines.html.

81. Cevik M, Marcus JL, Buckee C, Smith TC. Severe Acute Respiratory Syndrome Coronavirus 2 (SARS-CoV-2) Transmission Dynamics Should Inform Policy. Clin Infect Dis 2021; 73(Supplement_2): S170–S6.

82. James AE, Gulley T, Kothari A, Holder K, Garner K, Patil N. Performance of the BinaxNOW coronavirus disease 2019 (COVID-19) Antigen Card test relative to the severe acute respiratory coronavirus virus 2 (SARS-CoV-2) real-time reverse transcriptase polymerase chain reaction (rRT-PCR) assay among symptomatic and asymptomatic healthcare employees. Infect Control Hosp Epidemiol 2022; 43(1): 99–101.

83. Kahn R, Holmdahl I, Reddy S, Jernigan J, Mina MJ, Slayton RB. Mathematical Modeling to Inform Vaccination Strategies and Testing Approaches for Coronavirus Disease 2019 (COVID-19) in Nursing Homes. Clin Infect Dis 2022; 74(4): 597–603.

84. See I, Paul P, Slayton RB, et al. Modeling Effectiveness of Testing Strategies to Prevent Coronavirus Disease 2019 (COVID-19) in Nursing Homes—United States, 2020. Clin Infect Dis 2021; 73(3): e792–e8.

85. Goyal A, Reeves DB, Cardozo-Ojeda EF, Schiffer JT, Mayer BT. Viral load and contact heterogeneity predict SARS-CoV-2 transmission and super-spreading events. Elife 2021; 10.

86. Gomez J, Prieto J, Leon E, Rodríguez A. INFEKTA—An agent-based model for transmission of infectious diseases: The COVID-19 case in Bogotá, Colombia. PLoS One 2021; 16(2): e0245787.

87. Zipfel CM, Paul P, Gowler CD, et al. Modeling the Effectiveness of Healthcare Personnel Reactive Testing and Screening for the Severe Acute Respiratory Syndrome Coronavirus 2 (SARS-CoV-2) Omicron Variant Within Nursing Homes Clin Infect Dis 2022; 75(Supplement_2): S225–S30.

88. Jääskeläinen AE, Ahava MJ, Jokela P, et al. Evaluation of three rapid lateral flow antigen detection tests for the diagnosis of SARS-CoV-2 infection. J Clin Virol 2021; 137: 104785.

89. Goodall BL, LeBlanc JJ, Hatchette TF, Barrett L, Patriquin G. Investigating the Sensitivity of Nasal or Throat Swabs: Combination of Both Swabs Increases the Sensitivity of SARS-CoV-2 Rapid Antigen Tests. Microbiol Spectr 2022; 10(4): e0021722.

90. Ontario Health Canada. COVID-19 Rapid Antigen Tests: How to Collect a Sample. 2022. https://www.ontariohealth.ca/sites/ontariohealth/files/2022-02/COVID-19RapidAntigenTests-HowtoCollectaSample.pdf.

91. Reuters. Israel’s Health Ministry recommends throat swab for COVID-19 tests while U.S. FDA warns against. 2022. https://www.ctvnews.ca/health/coronavirus/israel-s-health-ministry-recommends-throat-swab-for-covid-19-tests-while-u-s-fda-warns-against-1.5734130.

## Supplementary Reference

1. Carter Alyssa M, Viloria Winnett A, Romano Anna E, Akana R, Shelby N, Ismagilov RF. Laboratory Evaluation Links Some False-Positive COVID-19 Antigen Test Results Observed in a Field Study to a Specific Lot of Test Strips. Open Forum Infectious Diseases 2023: doi: 10.1093/ofid/ofac701.

